# KRAS^G12D^-Specific Targeting with Engineered Exosomes Reprograms the Immune Microenvironment to Enable Efficacy of Immune Checkpoint Therapy in PDAC Patients

**DOI:** 10.1101/2025.03.03.25322827

**Authors:** Valerie S. LeBleu, Brandon G. Smaglo, Krishnan K. Mahadevan, Michelle L. Kirtley, Kathleen M. McAndrews, Mayela Mendt, Sujuan Yang, Ana S. Maldonado, Hikaru Sugimoto, Maria E. Salvatierra, Luisa M Solis Soto, Rick Finch, Mihai Gagea, Adam C. Fluty, Steve J. Ludtke, J Jack Lee, Abhinav K. Jain, Gauri Varadhachary, Rachna T Shroff, Anirban Maitra, Elizabeth Shpall, Raghu Kalluri, Shubham Pant

## Abstract

Oncogenic *KRAS* drives initiation and maintenance of pancreatic ductal adenocarcinoma (PDAC). Here, we show that engineered exosomes with Kras^G12D^ specific siRNA (iExoKras^G12D^) reveal impressive biodistribution in pancreas with negligible toxicity in preclinical studies in mice and Rhesus macaques. Clinical testing of iExoKras^G12D^ in the iEXPLORE (iExoKras^G12D^ in Pancreatic Cancer) Phase I study employed a classical 3+3 dose escalation design (Phase Ia), followed by an accelerated titration design (Phase Ib) (NCT03608631). Patients with advanced metastatic disease were enrolled after failure of multiple lines of therapy. iExoKras^G12D^ therapy was well-tolerated with no reported dose-limiting toxicity with some cases of stable disease response, and maximum tolerated infusion was not reached even at the highest dose. Downregulation of *KRAS^G12D^*DNA and suppression of phopho-Erk was documented with increased intratumoral in CD8^+^ T cell infiltration in patient samples upon treatment. The CD8^+^ T cell recruitment priming by iExoKras^G12D^ informed on potential efficacy of immune checkpoint therapy and lead to validation testing in preclinical PDAC models. Combination therapy of iExoKras^G12D^ and anti-CTLA-4 antibodies, but not anti-PD1, revealed robust anti-tumor efficacy via FAS mediated CD8^+^ T cell anti-tumor activity. This first-in-human, precision medicine clinical trial offers new insights into priming of immunotherapy by oncogenic Kras inhibitor and an opportunistic combination therapy for PDAC patients.

## Introduction

Pancreatic ductal adenocarcinoma (PDAC) diagnosis is near uniformly lethal, with a median survival of less than 12 months^1,2^, despite the availability of combination chemotherapy regimens. The pivotal driver oncogenic event in PDAC is mutant *KRAS*, implicated in ∼95% of tumors, with *KRAS^G12D^* found in half of PDAC cases with mutant *KRAS*^3^. Genetically engineered mice (GEM) modelling PDAC have demonstrated the prerequisite for mutant *Kras* in the initiation of PDAC, as well as the remarkable dependence of oncogenic Kras for cancer maintenance and extinction. In advanced stages of PDAC in mice, genetic extinction of mutant Kras resolves tumors and rescues mice from PDAC lethality^4^. We recently reported on the cancer cell expression of FAS death receptor and permissive FASL^+^CD8^+^ T cell-mediated anti-tumor response with Kras extinction in PDAC GEM^4^. Specifically, the suppression of oncogenic Kras in PDAC GEM relieved the epigenetic suppression of FAS death receptor on cancer cells, facilitating CD8^+^ T cell FASL-FAS mediated eradication of cancer cells^4^. Additionally, pharmacological inhibition of Kras^G12D^ with an allele-specific small molecule inhibitor (MRTX1133) also enabled CD8^+^ T cell targeting of cancer cells re-expressing FAS, in both GEM and patient-derived organoids^5^. Combination therapy with MRTX113 and CTLA-4 antibodies showed significant survival advantage compared to controls, supporting synergistic anti-tumor efficacy for oncogenic Kras targeting with immune checkpoint blockade (ICB) therapy^5^. We and others also reported on the role of oncogenic Kras in shaping the suppressive metabolic and immune tumor microenvironment of PDAC^6–8^ generating hypotheses for possible synergistic opportunities with immunotherapies. Collectively these studies offered insight on the failure of ICB in patients with PDAC – that the lack of clinical efficacy observed with ICB in patients with PDAC may be attributable to intrinsic and microenvironmental hurdles requiring specific combination therapy to sensitize PDAC to the therapeutic impact of ICB. A critical aspect in overcoming ICB ineffectiveness would include strategies of sustained and specific suppression of oncogenic Kras expression to generate lasting impact concurrent with ICB, while avoiding on-target, off-tumor collateral effects.

Although oncogenic Kras is recognized as a desirable therapeutic target in PDAC due to the near ubiquitous prevalence of “hotspot” mutations, it was dubbed the “undruggable” target due to its challenging chemistry^9^. More recently, groundbreaking oncogenic KRAS targeting with new direct small molecule inhibitors have made their way into clinical testing^10^, with long-awaited hope for enhancing PDAC survival. Despite the observed efficacy of monotherapy with available inhibitors, patients eventually develop resistance via pleiotropic mechanisms, and most experience at least some measure of treatment-related adverse events (most commonly, cutaneous and gastrointestinal)^11,12^. Keeping in mind the numerous challenges of targeting oncogenic Kras, we devised an alternative strategy that employed exosomes as a delivery vehicle for siRNA targeting. Exosomes or small extracellular vesicles (sEVs) are naturally produced lipid bilayer vesicles with the topography of their cell of origin, ranging in size from 40-200 nm, and with a regulated cargo that reflects cellular constituents^13^. Exosomes are a complex constituent of the systemic circulation and were demonstrated to likely play a role in inter-cellular communication^13,14^. The siRNA sequences used were previously reported to target oncogenic Kras rather than wild-type Kras, and we confirmed their specificity^15^. This effort was motivated by the observation that exogenously administered exosomes in mice accumulated in the pancreas and, when containing a Kras^G12D^ siRNA payload, significantly suppressed Kras signaling, resulting in increased survival of PDAC GEM with Kras^G12D^ mutation^16^. We coined these therapeutic exosomes as “iExoKras^G12D^”. We further demonstrated that iExoKras^G12D^ anti-tumor efficacy in mice relied on multiple aspects of exosomal biology, including their favored uptake in oncogenic Kras expressing cells via macropinocytosis^16^. Our studies also identified that the CD47 “don’t eat me” signal on iExoKras^G12D^ enabled superior systemic half-life compared to synthetic particles, with impact on efficacy of siRNA therapeutic payload in PDAC GEM^16^. Challenges for personalized therapy with exosomes included good manufacture practice (GMP) and large-scale production of a clinically defined product. We reported on the feasibility of iExoKras^G12D^ GMP production for human testing^17^, which led to approval for first in human clinical testing. Here, we tested the safety of iExoKras^G12D^ in murine and non-human primates (NHP) and provide a report. We also describe the results from the Phase I trial of iExoKras^G12D^ monotherapy in advanced PDAC. We designed a 3+3 clinical trial with escalading dose of iExoKras^G12D^ for metastatic PDAC with standard of care therapy failure (Phase Ia). Lack of dose-limiting toxicities (DLT) informed the follow up accelerated dosing strategy, again yielding no measurable toxicity (Phase Ib). We also report on studies with patient-derived biopsies that informed on potential synergistic combination of iExoKras^G12D^ with immune checkpoint blockade, specifically anti-CTLA-4, and subsequent murine studies validating this synergistic approach for Phase II trial design. Collectively, our preclinical studies, guided by the clinical trial, inform on a novel combination therapy of oncogenic Kras targeting with anti-CTLA-4 immune checkpoint blockade for robust anti-tumor response with potential to overcome resistance.

## Results

### GMP grade exosomes for siRNA delivery showed no toxicity in preclinical studies

The oncogenic Kras targeting approach we devised employed exosomes purified from the culture supernatant of bone marrow derived stromal cells from a healthy donor (**Figure 1a**). The purified exosomes were subjected to electroporation for the incorporation of siRNA with a sequence targeting KRAS^G12D^ (**Figure 1a**). The resulting exosomes were then processed for quality control (QC), which included size assessment by NanoSight^TM^ (**Supplementary** Figure 1A) and flow cytometry analyses for CD47 and putative exosomal marker CD63 (**Supplementary** Figure 1B**-C**). Following electroporation with siRNA, the exosomes were referred to as ‘iExoKras^G12D^’. Additional validation with exosomes marker, CD9, CD81 are also provided (**Supplementary** Figure 1C). The iExoKras^G12D^ were then quantified using microBCA assay to define the exosomal protein content used for dosing definition (*discussed below*). The siRNA content in the iExoKras^G12D^ equaled that of the exosomal protein content (1:1 by weight). We previously detailed the GMP, large-scale production of iExoKras^G12D^, including quality control assessment and preclinical testing showing anti-tumor effect with increased survival in PDAC GEM^17^. Scanning electron microscopy of iExoKras^G12D^ showed a distribution of exosomes (**Supplementary** Figure 1D) consistent with NanoSight^TM^ size distribution measurements. Cryogenic electron microscopy (Cryo-EM) revealed iExoKras^G12D^ lipid bilayer vesicles, some of which showed multiple vesiculation (**Figure 1B, Supplementary** Figure 1E), consistent with recent reporting^18^. Keeping in mind the clinical utility of GMP iExoKras^G12D^, we determined the toxicology profile of GMP iExoKras^G12D^ in mice and non-human primates (Rhesus macaques). Three cycles of iExoKras^G12D^ treatment administered intravenously, 3 doses per cycle for a total of 9 doses over the course of 6 weeks, was evaluated in healthy adult mice (**Figure 1C**). A comprehensive toxicology profile was established, evaluating diluent for iExoKras^G12D^ (PlasmaLyte) and increasing iExoKras^G12D^ doses (1X, 2X, and 4X, **Table 1**). Body weight showed no significant changes with any of the dose levels (**Figure 1D, Supplementary** Figure 2A), and liver and kidney weights were unchanged (**Supplementary** Figure 2B). Although a significant % change in spleen weight was registered for 1X iExoKras^G12D^, no significant changes were noted for higher doses (**Supplementary** Figure 2B). Comprehensive chemistry and hematology panels revealed no changes outside physiological ranges (**Supplementary** Figure 2C-D). When significant changes were observed when compared to the control group, those changes also remained within normal physiological range (**Supplementary** Figure 2C-D).

**Figure 1.**
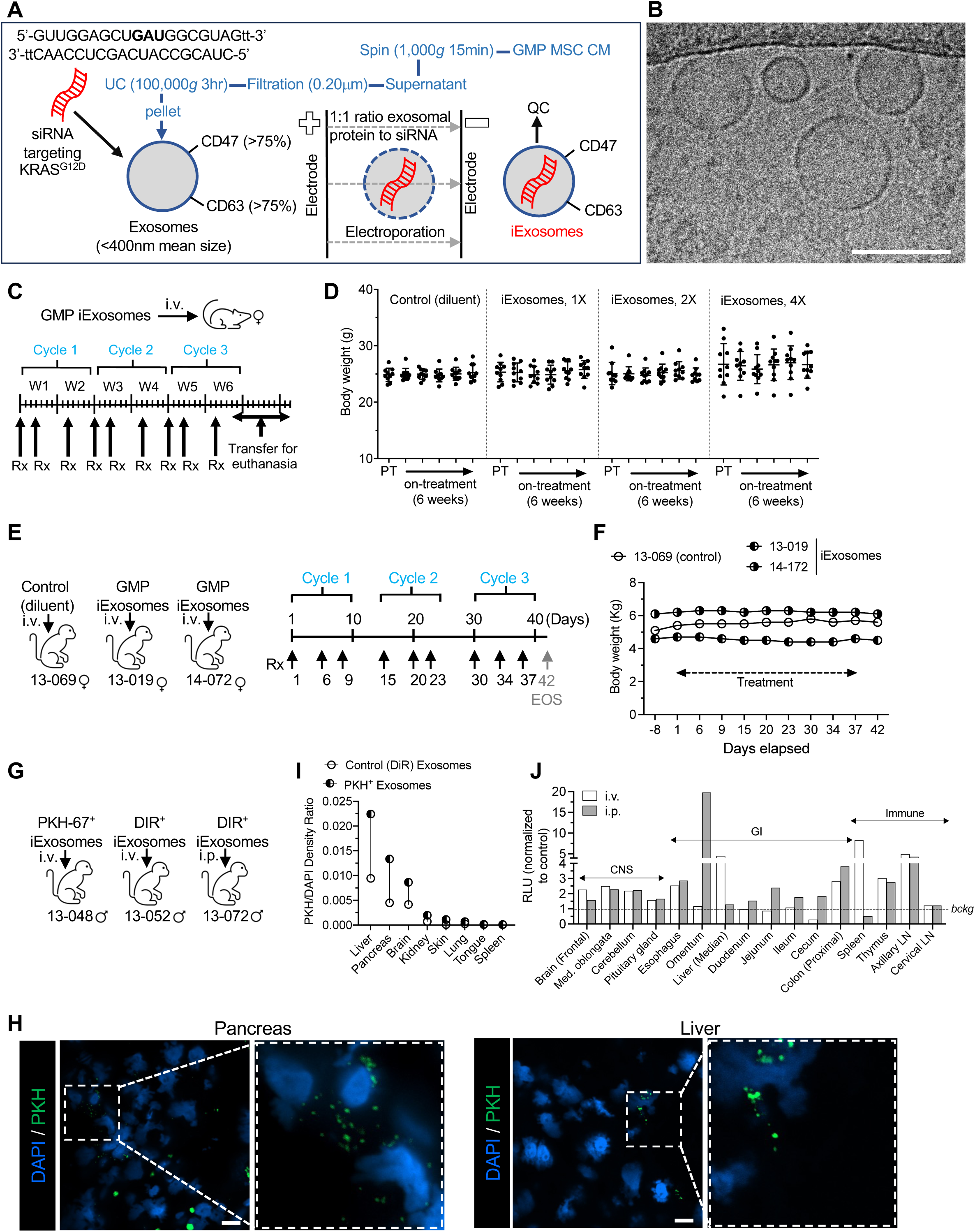
GMP iExoKras^G12D^ preclinical safety and biodistribution. **A**. Schematic representation of iExoKras^G12D^ GMP production. Conditioned media (CM) from mesenchymal stromal cells (MSC) cultured in GMP was centrifuged to remove debris, filtered through 0.2 um filter, and ultracentrifuged. The exosomes pellet was resuspended, subjected to electroporation to incorporate siRNA targeting KRAS^G12D^ using a 1:1 ratio (by weight) of exosomal protein and siRNA, and evaluated using quality control (QC) analysis for size distribution (mean size < 400 nm) and exosome surface detection of CD47 and CD63 by flow cytometry. **B**. Representative picture of cryo-electron microscopy of GMP iExoKras^G12D^. Scale bar: 100 nm. **C**. Schematic representation of murine toxicology study using intravenous (i.v.) GMP iExoKras^G12D^ treatment (Rx) for three consecutive cycles. Each cycle represents two weeks (W1: week 1; W2: week 2, etc.) and 3 iExoKras^G12D^ treatment on days 1, 4, 10. **D**. Body weight measurement over time for the listed groups of mice, PT: pretreatment. 1X, 2X, and 4X exosomes reflect dosing corresponding to clinical trial dosing, see also **Table 1**, n=10 mice per group. **E**. Schematic representation of NHP toxicology study using intravenous (i.v.) GMP iExoKras^G12D^ treatment (Rx) for three consecutive cycles. Each cycle represents two weeks (W1: week 1; W2: week 2, etc.) and iExoKras^G12D^ treatments on the listed days, distributing 3 iExoKras^G12D^ treatments over the course of 2 weeks. **F**. Body weight measurement over time for the listed NHP. For dosing details, see **Table 1**. **G**. Schematic representation of NHP biodistribution study using intravenous (i.v.) or intraperitoneal (i.p.) GMP iExoKras^G12D^ that was labeled with PKH-67 or DiR dye. **H-I** Representative fluorescence images (H) of pancreas and liver sections from NHP following PKH-67^+^ iExoKras^G12D^ treatment and associated quantification (I) of the density of the ratio of PKH67 level over nuclear staining (DAPI) in the listed organs. Scale bar: 10μm. **J**. Normalized relative luminescence unit (RLU) in the listed organs for the detection of DiR^+^ label following i.v. or i.p. DiR^+^ iExoKras^G12D^ treatment. CNS: central nervous system; GI: gastrointestinal; Med. oblongata: medulla oblongata; LN: lymph node.

**Table 1.**

|  | Murine (average BW = 25g) |  |  | NHP (average BW = 6 kg) |  |  | Human (average BW = 70kg) |  |  |  |  |
| --- | --- | --- | --- | --- | --- | --- | --- | --- | --- | --- | --- |
|  | iExo (1:1 Exo:siRNA, exp. as Exo protein) | siRNA or Exo |  | iExo (1:1 Exo:siRNA, exp. as Exo protein) | siRNA or Exo |  | iExo (1:1 Exo:siRNA, exp. as Exo protein) | siRNA or Exo |  |  |  |
|  | mg/kg | mg/kg | mg per 25g BW | mg/kg | mg/kg | mg per 6kg BW | mg/kg | mg/kg | mg per 70kg | Clinical Trial Dose Levels |  |
| Preclinical anti-tumor studies (i.p.) | 0.04 | 0.04 | 0.001 | 0.0100 | 0.0100 |  | 0.00324 | 0.00324 |  | L1-L2* | *Clinical Trial equivalent |
| NHP Imaging study (i.v. & i.p.) |  |  |  | 0.0110 | 0.0110 | 0.070 |  |  |  |  |  |
| NHP tox study (i.v) |  |  |  | 0.0021 | 0.0021 | 0.150 |  |  |  | L1* | *Clinical Trial equivalent |
| Murine tox study (i.v.) | 0.026 | 0.026 | 0.66 |  |  |  | 0.0021 | 0.0021 | 0.150 | L1 | Clinical Trial (i.v.) |
|  | 0.053 | 0.053 | 1.32 |  |  |  | 0.0043 | 0.0043 | 0.300 | L2 |  |
|  | 0.106 | 0.106 | 2.65 |  |  |  | 0.0086 | 0.0086 | 0.600 | L3 |  |
|  |  |  |  |  |  |  | 0.0172 | 0.0172 | 1.200 | L4 |  |
|  |  |  |  |  |  |  | 0.0344 | 0.0344 | 2.400 | L5 |  |
|  |  |  |  |  |  |  | 0.0688 | 0.0688 | 4.800 | L6 |  |

Toxicology studies were also evaluated in non-human primates (NHP), following a similar dosing strategy as planned for clinical testing (**Figure 1E**, **Table 1**). Three Rhesus macaques were treated with 9 doses of GMP iExoKras^G12D^ over the course of 6 weeks (**Figure 1E**, **Table 1**). Body weight measurements were unchanged (**Figure 1F, Supplementary** Figure 3A). Organ weight, chemistry and hematological studies were minimally or insignificantly altered (**Supplementary** Figure 3B-C). Gross examination was unremarkable, and urine analysis and a detailed anatomic pathology evaluation revealed no lesions or histopathological changes. Furthermore, we investigated the biodistribution of exogenously administered GMP iExoKras^G12D^ in Rhesus macaques 24 hours following intravenous (i.v.) or intraperitoneal (i.p.) injections (**Figure 1G**, **Table 1**). In this experimental design, PKH-67^+^ labeled iExoKras^G12D^-infused NHP (i.v.) served as control for DiR^+^ labeled iExoKras^G12D^ (and in reciprocal). PKH-67^+^ exosomes were visualized in pancreas, liver, and brain tissue sections with confocal and high-resolution fluorescence microcopy, with minimal positive PKH-67^+^ exosomes noted in kidney, lung, skin, spleen, and tongue (**Figure 1I-J, Supplementary** Figures 4-5). Histopathological examination of pancreas, liver, and brain was unremarkable (**Supplementary** Figure 6A). Whole tissue bioluminescence capture of DiR^+^ signal in NHP administered with DiR^+^ labeled iExoKras^G12D^ i.v. vs i.p. showed positive signal in CNS tissues, gastrointestinal tract, and immune organs including lymph nodes (**Figure 1J**), with minimal positive foci noted in kidney, lung, skin, spleen, and tongue (**Supplementary** Figure 6B). While liver capture of DiR^+^ signal was greater following i.v. administration of iExoKras^G12D^, omentum capture of DiR^+^ signal was prominent following i.p. administration (**Figure 1J**). In this experimental setting however, high background autofluorescence of certain control tissues, including occipital and parietal lobes of the cortical brain, pancreas, and prostate, prevented accurate capture of DiR^+^ signal (**Supplementary** Figure 6B). Quantification for *KRAS^G12D^* siRNA by digital PCR analysis showed detection of siRNA from iExoKras^G12D^ in liver, pancreas, and bone marrow (**Supplementary** Figure 7). Levels of siRNA were higher in the liver when iExoKras^G12D^ were administered i.v., with the highest level of siRNA captured in the pancreas of NHP following i.v. iExoKras^G12D^, albeit RNA degradation prevented comparative analysis of siRNA in one i.v. treated NHP pancreas (**Supplementary** Figure 7A-B). Taken together, the results support iExoKras^G12D^ administration into mice and Rhesus macaques is safe, with a biodistribution indicative to iExoKras^G12D^ accumulation in pancreas and liver.

### Systemic iExoKras^G12D^ treatment showed no measurable toxicity in dose escalation clinical trial

Clinical testing of iExoKras^G12D^ was initiated with a 3+3 dose escalation (Phase Ia) and enrolled 9 evaluable patients with metastatic pancreatic cancer at three dose levels (**Figure 2A**). The three dose levels were equivalent to dose levels tested in toxicology studies in mice and NHP (**Table 1**). iExoKras^G12D^ dosing was defined using exosomal protein level (which contained an equivalent weight of siRNA payload [**Table 1]**). Patient eligibility included histologically confirmed metastatic PDAC with *KRAS^G12D^* mutation (confirmed in tissue or blood), progression of disease on one or more line of systemic therapy in the metastatic setting (or stable disease with at least 4 months of chemotherapy with cytotoxic therapy), and Eastern Cooperative Oncology Group (ECOG) performance status of 0 and 1. Consented patients were at least 18 years of age, with negative pregnancy serum testing for women of childbearing potential and adequate contraception for males. Exclusion criteria included pregnancy, concurrent severe or uncontrolled stable angina, myocardial infarction within 6 months, symptomatic arrythmia, uncontrolled diabetes, serious active infection, central nervous system disease (with exception of treated brain metastasis). In Phase Ia, patients received 3 doses of iExoKras^G12D^ over the course of 2 weeks (1 cycle) for three consecutive cycles (**Figure 2B**). Response was assessed by imaging using Response Evaluation Criteria in Solid Tumors (RECIST) 1.1, utilizing the Quantitative Imaging Analysis Core (QIAC), a centralized platform for diagnostic imaging at the MD Anderson Cancer Center. Dose-limiting toxicity grading followed the National Cancer Institute Common Terminology Criteria for Adverse Events (NCI CTCAE, Version 4.0). Six males and three females, with a range of 49 to 75 years of age were enrolled (**Figure 2C**). All patients received multiple lines of prior systemic therapies and showed time from diagnosis to cancer related death ranging from 1 year (or less) to 7 years (**Figure 2C**). *KRAS^G12D^* mutation status was confirmed along with additional mutations (**Figure 2C**). Three patients, one each at dose level 1, 2, and 3, showed stable disease after 3 cycles and were enrolled in additional iExoKras^G12D^ treatment, per protocol (**Figure 2B-C**). One patient, enrolled in the dose 3 level, showed symptomatic progression of disease after receiving just one iExoKras^G12D^ infusion (less than 1 cycle) and was excluded from further analyses and replaced, given that rapid progression precluded continuation of 3 consecutive cycles (**Figure 2C**). All but one patient that completed at least 3 cycles of iExoKras^G12D^ therapy, including those with initial stable disease staging on iExoKras^G12D^, eventually progressed with new lesions and/or target lesions (**Figure 2C**). S3D1 showed stable disease after completion of 6 cycles of iExoKras^G12D^ and was found subsequently to have progression of disease 3 months after completing the iExoKras^G12D^ trial. No related adverse events were noted for any patients at the dose levels tested in the 3+3 Phase Ia trial (**Figure 2C**). Given lack of toxicity in all dose levels tested, a Phase Ib with accelerated dose titration was designed, with 3 additional patients enrolled at increased dose level for each patient (**Figure 2A-B**). To ease patient infusion schedule, the accelerated titration Phase Ib study was revised with weekly administration, for a total of 6 infusions over the course of 6 weeks (**Figure 2B**). Tissue biopsies were captured pre- and post-iExoKras^G12D^ treatment (**Figure 2B**). Two females and one male in an age range of 48 to 76 years and with confirmed *KRAS^G12D^* status were treated with increasing levels of iExoKras^G12D^ (**Figure 3A**). Timeline of disease progression, prior therapy, time of biopsy for mutation analysis, and visual display of target lesion monitoring indicated iExoKras^G12D^ therapy represented a short window of time compared to prior treatments (**Figure 3B**). All three patients showed stable disease and enrolled for additional iExoKras^G12D^ dosing before showing progression and growth of target lesions (**Figure 3A-B**). Patient disease progression in this accelerated titration Phase Ib trial was between 1 (or less) to 2 years from cancer diagnosis to cancer related death (**Figure 3A**). Though patients demonstrated an ECOG status of 0 and 1 prior to enrollment, Phase Ib iExoKras^G12D^ treatment was administered to patients with 1-to-2 years disease progression from diagnosis to cancer related death (**Figure 3B**), and at 4-to-8-month range of time prior to cancer related death. In the Phase Ia/b clinical trial, iExoKras^G12D^ was well tolerated with no treatment-related adverse events reported and the maximum tolerated dose not reached (**Figure 3A**), thereby demonstrating a favorable safety profile. The highest dose level tested was 4.8 mg of iExoKras^G12D^ containing 4.8 mg of siRNA per dose, with 6 consecutive doses totaling 28.8 mg of iExoKras^G12D^ over the course of 12 weeks.

**Figure 2.**
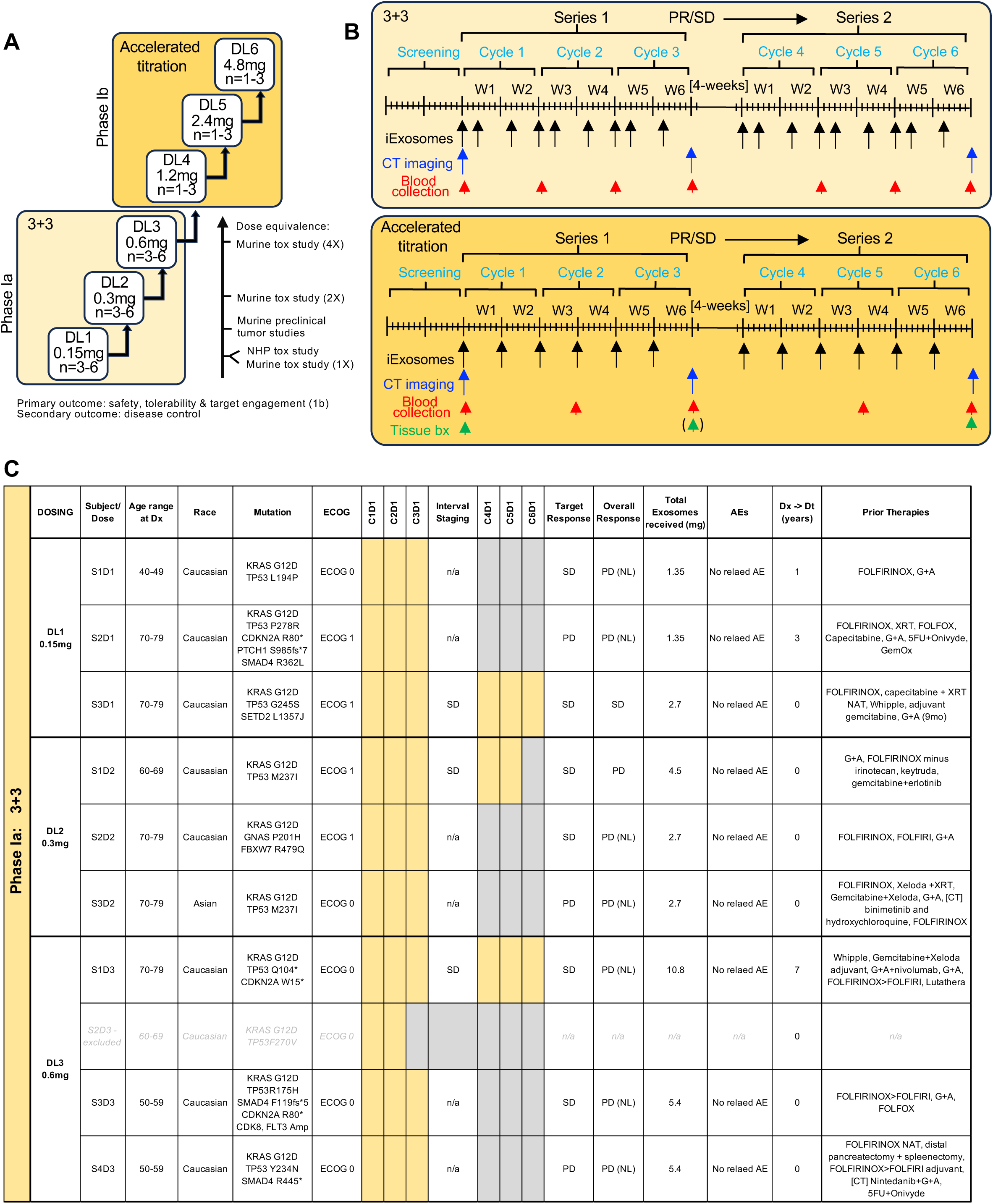
GMP iExoKras^G12D^ clinical trial design. **A.** Schematic representation of dose escalation design. DL1: dose level 1; DL 2: dose level 2; etc. **B.** Schematic representation of dosing schedule. PR/SD: partial response/stable disease; bx: biopsy **C.** Patient demographic, defining characteristics (including prior therapies), treatment cycle completion, response rate, total iExoKras^G12D^ received (expressed as the cumulative sum of exosomal proteins), and adverse events (AEs). Dosing cycles in yellow: dosing completed. Dosing cycles in grey: no treatment. C1D1: Cycle 1 Day1; C2D1: Cycle 2 Day 1; etc. M: male; F: female; SD: stable disease; PD: progressive disease; NL: new lesion; Dx -> Dt: time in year from diagnosis (Dx) to cancer related death (Dt). G+A: gemcitabine + Abraxane; XRT: radiation therapy; GemOx: gemcitabine + oxaliplatin; [CT]: other clinical trial; NAT: neoadjuvant therapy; 5FU: 5-fluorouracil; n/a: not applicable.

**Figure 3.**
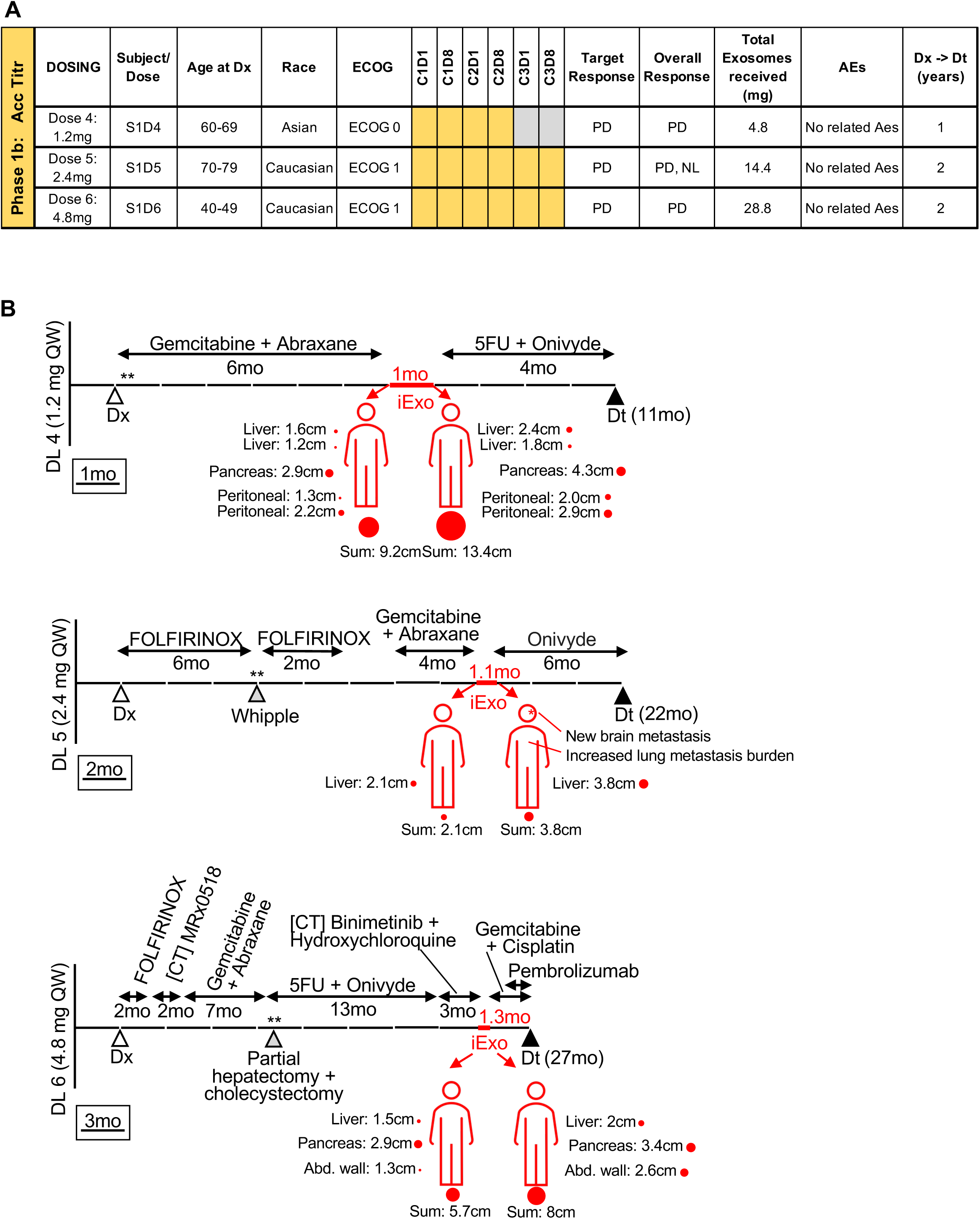
GMP iExoKras^G12D^ in accelerated titration dosing design. **A.** Patient demographic, defining characteristics, treatment cycle completion, response rate, total iExoKras^G12D^ received (expressed as the cumulative sum of exosomal proteins), and adverse events (AEs). Dosing cycles in yellow: dosing completed. Dosing cycles in grey: no treatment. M: male; F: female; SD: stable disease; PD: progressive disease; NL: new lesion; Dx ® Dt: time in year from diagnosis (Dx) to cancer related death (Dt). n/a: not applicable. **B.** Schematic representation of treatment timeline for patients in DL 4: dose level 4, DL 5: dose level 5, and DL5, and DL6: dose level 6, depicting prior therapy, timeframe for iExoKras^G12D^ therapy, and target lesion progression; scale bar listed. C1D1: Cycle 1 Day1; C2D1: Cycle 2 Day 1; etc. QW: weekly; Dx: diagnosis; mo: month; Dt: death; [CT]: clinical trial; **: mutation analysis.

### Systemic iExoKras^G12D^ treatment showed target engagement and remodeled tumor immune microenvironment

We previously showed GMP iExoKras^G12D^ targeting Kras^G12D^ in PDAC GEM synergized with gemcitabine chemotherapy to limit tumor growth and increase survival^17^. We first conducted preclinical studies to determine if serial liquid biopsies for circulating *Kras^G12D^* DNA would serve as a surrogate for tumor response. Following orthotopic cancer cell injection, mice were monitored for tumor burden by IVIS imaging. Prior to treatment start (pre-tx), mice were bled (**Figure 4A**). The level of circulating *Kras^G12D^* DNA was compared in blood collected prior to iExoKras^G12D^ treatment and 9 days after treatment, with treatment groups including gemcitabine, GMP iExoKras^G12D^, or a combination of iExoKras^G12D^ and gemcitabine treated mice, and with control group including mice treated with diluent or control, non-electroporated exosomes (CE: control exosomes)^17^. As cancer progressed in control groups, the relative level of *Kras^G12D^*DNA (expressed as a percent of *Kras^G12D^* copies over total *Kras* copies) increased with tumor burden (**Figure 4B**). Suppression of tumor growth, with gemcitabine, iExoKras^G12D^, or combination thereof, tracked with lower relative levels of circulating *Kras^G12D^* (**Figure 4B**). iExoKras^G12D^ therapy and combination therapy with iExoKras^G12D^ showed lowest relative levels of circulating *Kras^G12D^* (**Figure 4B**), congruent with target specific engagement and anti-tumor response^17^. Mimicking the preclinical data, assessment for relative levels of circulating *Kras^G12D^* in patients treated with iExoKras^G12D^ in the Phase I trial demonstrated reduced mutant allele fraction in several patients, including S2D2, S1D3, S4D3, as well as S1D5 (**Figure 4C-D**).

**Figure 4.**
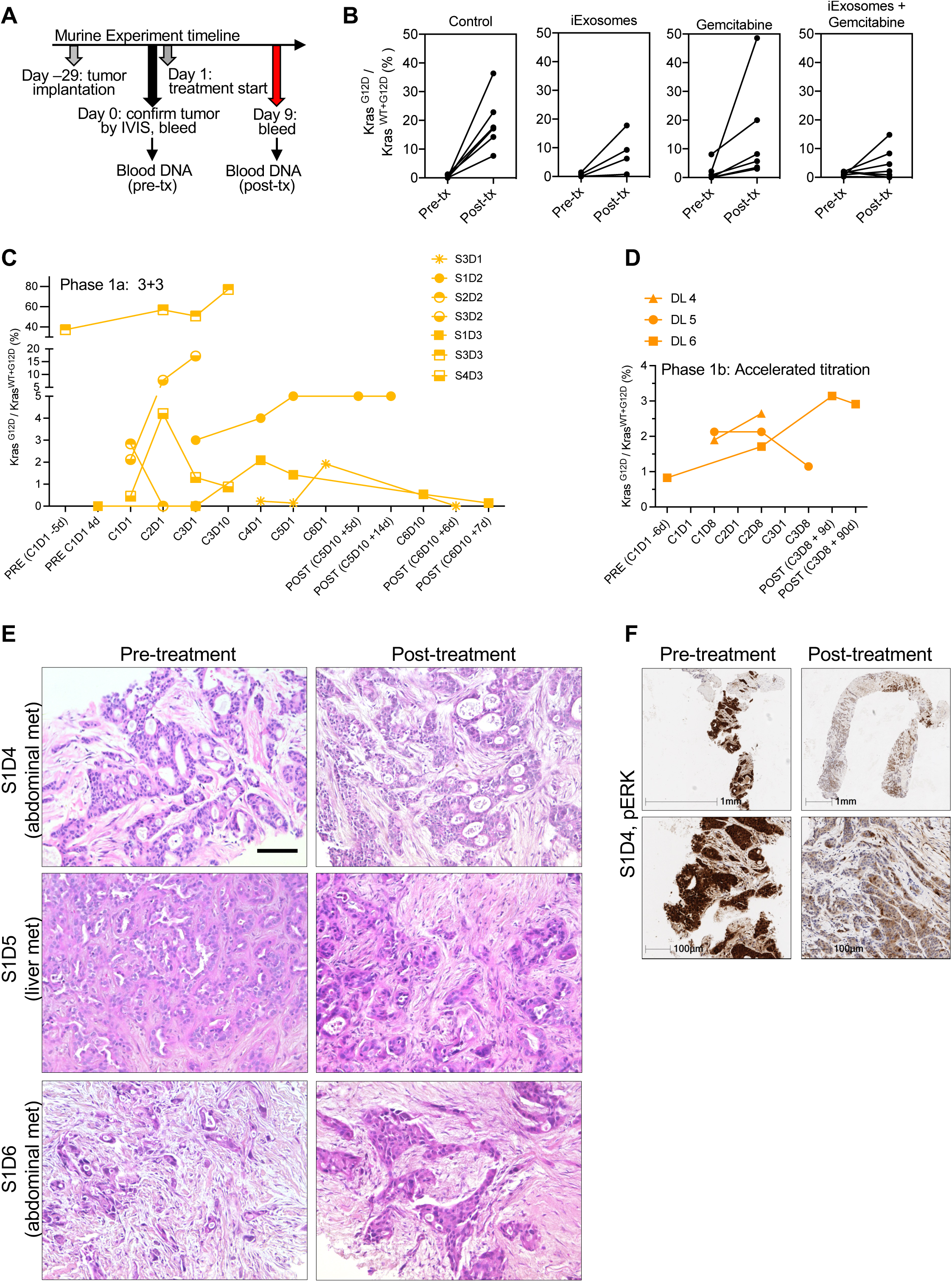
cfDNA and biopsy analyses. **A**. Schematic representation of murine experimental timeline for tumor inoculation, IVIS imaging of tumor burden, and blood collection. **B**. Relative level of *Kras^G12D^* cfDNA (expressed as a percent of *Kras^G12D^* copies over total *Kras* copies) in pre-treatment (pre-tx) and post-treatment (post-tx) with iExoKras^G12D^ and/or gemcitabine. Each line represents a distinct mouse per the listed experimental group: Control n=6; iExoKras^G12D^ n=4; gemcitabine n=6; iExoKras^G12D^ + gemcitabine n=7 mice. **C-D**. Relative level of *KRAS^G12D^* cfDNA in the listed patients over time in the 3+3 (C) and accelerated dose titration (D) studies. PRE: pre-treatment; POST: post-treatment, with cycle definition at each time point evaluated. **E**. Representative H&E-stained tumor section from metastases in the abdominal wall (abdomen) or liver pre- and post-treatment with iExoKras^G12D^. Scale bar upper panel: 200 μm, lower panel 100 μm. **F.** Representative pERK-stained tumor section from metastases in the abdominal wall of S1D4, scale bar listed.

Tissue biopsies pre- and post-treatment were captured in the accelerated titration Phase Ib trial, and sections comprising of cancer cells and stromal cells, indicated by H&E staining (**Figure 4E**), were selected for further immunolabeling studies. Down regulation in phosphorylated ERK, a downstream effectors indicative of active RAS signaling (**Figure 4F**), decrease number of PanCK^+^ cancer cells, and stable or relative increase in αSMA^+^ stromal cells noted post-treatment (**Figure 5A-B**). Analysis of T cell infiltrates revealed an increase in intratumoral CD8^+^ T cells, CD4^+^ Foxp3^+^ Tregs and CD4^+^ Foxp3^-^ cells post-treatment compared to pre-treatment (**Figure 5C-D, Supplementary** Fig. 8A). Taken together, these results highlight the consistent target engagement of iExoKras^G12D^ in human PDAC with remodeling of the tumor immune microenvironment that likely favors to anti-tumor response, similar to that observed in murine PDAC treated with iExoKras^G12D4,5^.

**Figure 5.**
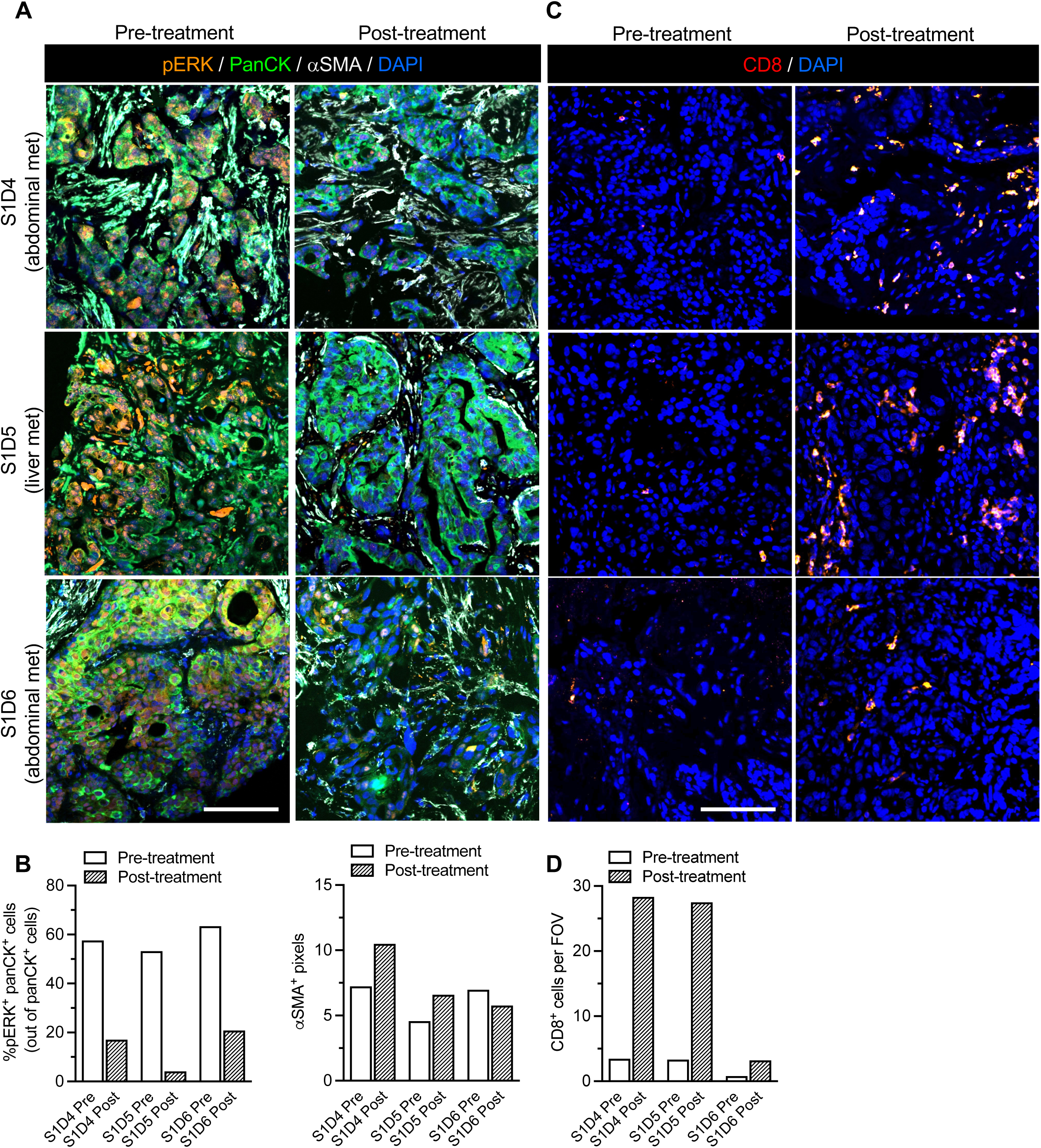
Target engagement with iExoKras^G12D^ treatment. **A-B**. Representative images of immunolabeling for pERK, PanCK, αSMA (**A**), with quantification of percentage of pERK^+^ panCK^+^ cancer cells (out of panCK^+^ cells) and αSMA^+^ pixels (**B**) in paired biopsies. **C-D.** Representative images (**C**), and quantification (**D**) of CD8^+^ T cells infiltrates in paired biopsies. Data are presented as mean. Scale bar:100 μm.

### CD8^+^ T cell recruitment and CD4^+^ T cells or Tregs depletion favored iExoKras^G12D^ therapy in preclinical models of PDAC

Informed the clinical data therein, we tested the efficacy of iExoKras^G12D^ therapy in combination with immune checkpoint blockade to enhance overall anti-tumor response. GMP-iExoKras^G12D^ downregulated *KRAS^G12D^* transcript in Panc-1 cells (**Supplementary** Figure 8B). Given the increase in intratumoral CD8^+^ T effectors and CD4^+^ Foxp3^+^ Tregs in clinical trial biopsies following iExoKras^G12D^ therapy, we next tested the functional contribution of these cells in PDAC following Kras^G12D^ inhibition with iExoKras^G12D^. T cell infiltration following Kras^G12D^ inhibition in established tumors of the autochthonous KTC (*P48-Cre, LSL-Kras^G12D/+^, Tgfbr2^F/F^*)^16^ PDAC GEM was measured (**Figure 6A**). Analysis of the intratumoral T cell populations revealed an increase in infiltration of CD4^+^ and CD8^+^ T cells in the PDAC tumor microenvironment (TME) following iExoKras^G12D^ treatment (**Figure 6B-C**), consistent with the findings in clinical samples (**Figure 5E-I**). Similar results were also noted in a second orthotopic KPC-689 PDAC model treated with iExoKras^G12D^ ^16^ (**Supplementary** Figure 9A), resulting in increased tumor infiltrating CD4^+^ and CD8^+^ T cells with iExoKras^G12D^ treatment (**Supplementary** Figure 9B). To determine the functional contribution of the T cell infiltrates following Kras^G12D^ inhibition, we crossed the KPC (*Pdx1-Cre, LSL-Kras^G12D/+^, Trp53^R172H/+^*) GEM with CD4^−/−^ or CD8^−/−^ mice and generated KPC mice depleted of CD4^+^ T cells or CD8^+^ T cells, respectively. KPC, KPC CD4^−/−^ and KPC CD8^−/−^ mice were treated with iExoKras^G12D^ and tumor burden was monitored by serial MRI (**Supplementary** Figure 9C-D). The control (no iExoKrasG12D) KPC, KPC CD4^−/−^ and KPC CD8^−/−^ mice showed similar tumor progression and survival kinetics (**Supplementary** Figure 9D**-E**). KPC mice treated with iExoKras^G12D^ demonstrated prolonged survival compared to control KPC mice (**Supplementary** Figure 9E), consistent with our prior findings^16^. KPC CD4^−/−^ mice treated with iExoKras^G12D^ demonstrated robust tumor growth inhibition by MRI and prolonged survival compared to control KPC CD4^−/−^ mice and iExoKras^G12D^ treated KPC mice (**Supplementary** Figure 9D-E). In contrast, KPC CD8^−/−^ iExoKras^G12D^ treated mice showed no survival benefit compared to the control KPC CD8^−/−^ mice (**Supplementary** Figure 9D-E), indicating that CD8^+^ T cells are required for Kras^G12D^ inhibition mediated suppression of PDAC^5^. Taken together, our data demonstrate that depletion of CD4^+^ T cells/T regs prime Kras^G12D^ inhibition and effector CD8^+^ T cells are required for PDAC control with iExoKras^G12D^ therapy.

**Figure 6.**
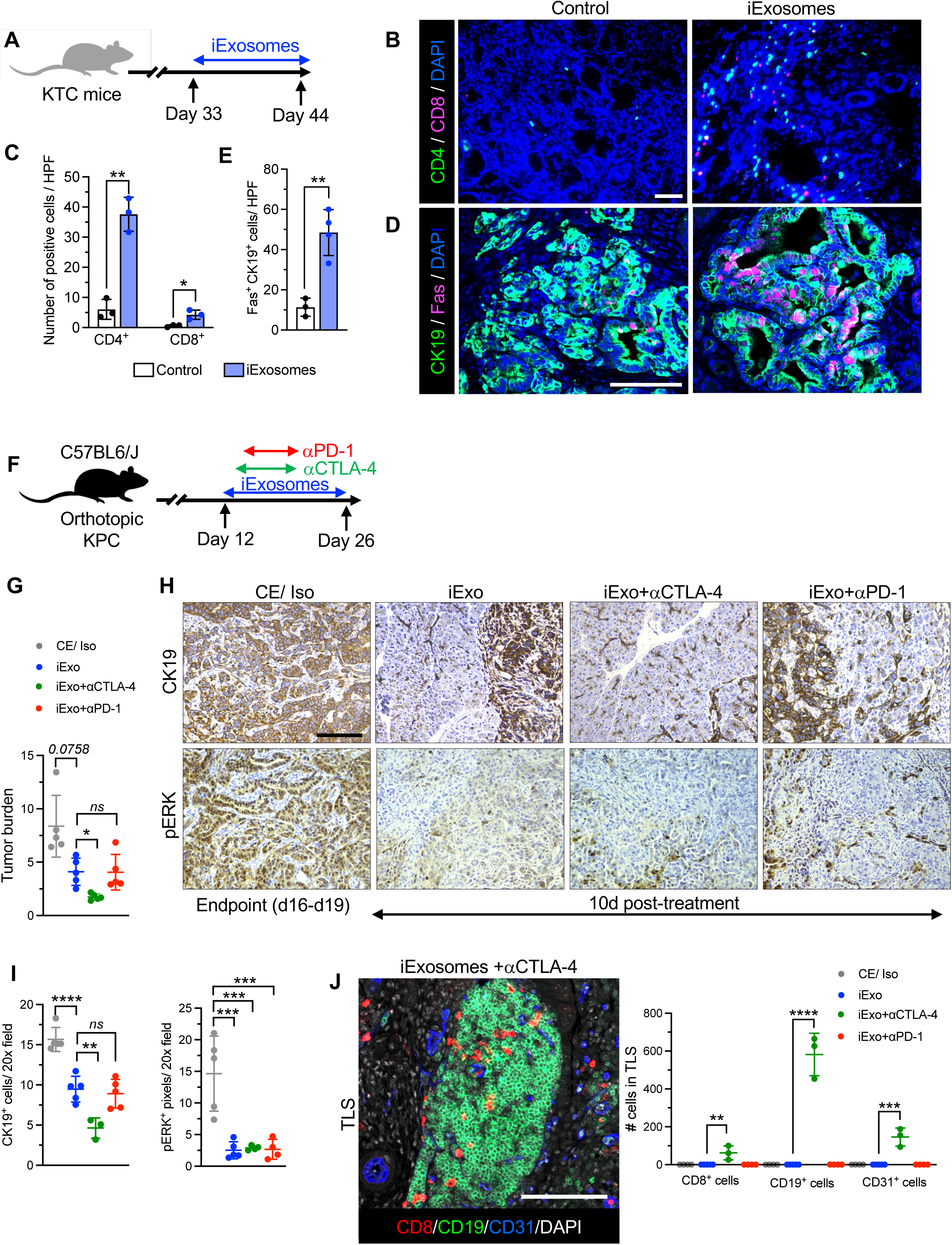
Synergy of iExoKras^G12D^ with anti-CTLA-4 for tumor suppression in preclinical studies. **A.** Schematic of experimental timeline using KTC GEM. iExoKras^G12D^ treatment was initiated at day 33 of age until day 44. **B-C**. Representative images (B) and respective quantification (C) of CD4^+^ T cells (CD4), and CD8^+^ T cells (CD8) by immunolabeling in control and iExoKras^G12D^ treated KTC mice, n=3 mice per group. **D-E**. Representative images (D) and respective quantification (E) of CK19^+^ and Fas^+^ cells by immunolabeling in control and iExoKras^G12D^ treated mice, n=3 mice per group. **F**. Schematic of experimental timeline using orthotopic PKC mice. Treatment was initiated at day 12 post tumor implantation and mice euthanized 2 weeks post-treatment (day 26). **G**. Tumor burden ((Pancreas or tumor weights /body weight) *100) of PKC mice treated with iExoKras^G12D^, control exosomes (CE), Isotypes control (Iso), αCTLA-4 and αPD-1, n=5 mice per group. **H-I**. Representative images (H), and quantification (I) of CK19 and pERK immunostainings in the indicated groups, n=3-5 mice per group. **J**. Representative image and respective quantification of CD8, CD19, CD31, and nuclear staining (DAPI) in tertiary lymphoid structure (TLS) in the tumors of mice in the indicated groups, n=3-5 mice per group. Scale bar: 100 μm. Data are presented as mean +/– SD. Significance was determined by unpaired t-test (B, C) or one-way ANOVA with Dunnett’s multiple comparisons test (E, G). αCTLA-4, anti-CTLA-4; αPD-1, anti-PD-1.

### iExoKras^G12D^ efficacy relies on Kras^G12D^ primed epigenetic regulation of Fas for Fas-FasL anti-tumor response by CD8^+^ T cells

*Fas* expression in pancreatic cancer cells following *Kras^G12D^*genetic suppression enables interaction with *FasL*-expressing CD8^+^ T cells and anti-tumor response^4,5^. CK19^+^ pancreatic cancer cells showed an increase in FAS expression (FAS^+^ CK19^+^) following iExoKras^G12D^ treatment in KTC GEM (**Figure 6D-E**). *In vitro*, KPC-689 cells treated with iExoKras^G12D^ demonstrated upregulation of expression in *Fas* and the CD8^+^ T cell recruiter cytokine, *IL-15* (**Supplementary** Figure 9F). Mechanistically, the regulation of *Fas* by Kras^G12D^ inhibition in pancreatic cancer cells included epigenetic remodeling, implicating methylation of the histones and recruitment of transcriptional repressor at the promoter site^4,19,20^. Chromatin immunoprecipitation (ChIP analysis) at the *Fas* promoter or transcriptional start site revealed that *Kras^G12D^* expression recruited DNA methyl transferase DNMT1 and histone methyl transferase EZH2, and iExoKras^G12D^ treatment inhibited DNMT1 and EZH2 recruitment (**Supplementary** Figure 9G). EZH2 promotes trimethylation of histone H3 at the K27 site^21^ and iExoKras^G12D^ treatment downregulated H3K27me3 and upregulated acetylation of H3K27 within the Fas promoter (**Supplementary** Figure 9G). Collectively, our data indicate that iExoKras^G12D^ treatment relieved Kras^G12D^ mediated epigenetic suppression of *Fas* in pancreatic cancer cells, enabling permissive immune TME priming to anti-tumor response via CD8^+^ T cell FASL-FAS mediated eradication of cancer cells.

### Clinical trial with iExoKras^G12D^ informed on combination therapy with immune checkpoint blockade

Based on the data collected, we next tested the efficacy of iExoKras^G12D^ therapy in combination with ICB to enhance anti-tumor response. Mice with orthotopic PDAC tumors (utilizing syngeneic cells derived from *P48Cre, LSL-Kras^G12D/+^, Trp53^F/+^* GEM) were treated with iExoKras^G12D^ and ICB (**Figure 6F**). Specifically, iExoKras^G12D^ treatment was combined with either anti-CTLA-4 or anti-PD-1 checkpoint immunotherapies (**Figure 6F**). Age matched analysis of PDAC tissues revealed a decrease in tumor burden with iExoKras^G12D^ therapy, with further decrease in tumor burden following anti-CTLA-4 combination but not with anti-PD-1 combination (**Figure 6G**). Immunolabeling of tumor sections revealed downregulation of pERK in all groups that received iExoKras^G12D^ (**Figure 6H-I**). Analysis of CK19^+^ cells (used here as surrogate for quantification of pancreatic cancer cells) revealed a decrease in CK19^+^ cancer cells following iExoKras^G12D^, with further decrease in mice treated with anti-CTLA-4 and iExoKras^G12D^ combination (**Figure 6H-I**). Histological analysis of PDAC tissues demonstrated an increase in the presence of tertiary lymphoid structures (TLS) in mice treated with iExoKras^G12D^ (iExo) + anti-CTLA-4 with 100% penetrance (**Supplementary** Figure 10A-B). The presence of TLS, characterized by formation of new lymphoid structures including B cells, CD8^+^ T cells, a newly developing vasculature with endothelial venules and antigen presenting cells^22–24^, is frequently associated with favorable responses to immunotherapy in the context of cancers such as melanoma, non-small cell lung carcinomas and better prognosis^23,24^. We performed TSA multiplex immunostaining analysis confirming the presence of CD8^+^ T cells, B cells, and endothelial cells in TLS accumulating in tumors of mice treated with iExoKras^G12D^ + anti-CTLA-4 (**Figure 6J**). Collectively these results support the superior anti-tumor response of iExoKras^G12D^ when combined specifically with anti-CTLA-4 therapy.

## Discussion

Our study demonstrates the safety of iExoKras^G12D^ as personalized medicine in patients with advanced PDAC. To our knowledge, this is the first-in-human study using exosomes from an allogenic source of bone marrow derived mesenchymal stromal cells that were engineered to encapsulate a siRNA therapeutic payload (iExoKras^G12D^) in a personalized medicine setting. Here we report on the lack of adverse events or toxicity of iExoKras^G12D^ in mice, NHP, and in PDAC patients. We also demonstrated the localization of exogenously administered exosomes to the liver and pancreas of NHP, as well as the target engagement of iExoKras^G12D^ in human PDAC. The target engagement results support the specificity of this approach to target oncogenes that are known drivers of cancer initiation and progression but for which pharmacological approaches are lacking.

Increase in dosing of systemic iExoKras^G12D^ in this Phase Ia/b clinical trial did not result in any adverse event and the maximum tolerated dose was not reached, suggesting iExoKras^G12D^ dosages may be further increased beyond the 28.8 mg/6weeks dose tested here to reach clinical efficacy in the planned Phase II studies. The safety profile of the iExoKras^G12D^ compared to other nanolipid-based carriers of siRNA or other molecular therapeutic cargo may reflect the intrinsic properties of exosomes^13^. Specifically, their natural lipid composition and surface protein expression reflect, at least in part, that of their cell of origin, minimizing their immunogenicity. Despite their allogenic source, no adverse events, including immunological reactions, were observed. The immune evasion of allogeneic and heterologous exosomes may rely on their surface expression of the CD47 don’t-eat-me signal, as previously reported^16^. The safety profile of iExoKras^G12D^ also lends itself to testing in combination with chemotherapy and immunotherapy. We previously reported on the synergistic impact of iExoKras^G12D^ in combination of gemcitabine in mice^17^.

We previously reported on the influx of intratumoral T cells following oncogenic Kras extinction and antitumor response mediated by re-expression of Fas on cancer cells, yielding anti-tumor response via its interaction with CD8^+^ T cells FasL^4,5^. Here we demonstrated influx of intratumoral CD8^+^ T cells in human biopsies following iExoKras^G12D^ therapy. Collectively, our data support not only target engagement by iExoKras^G12D^, but also biological response with remodeling of the immune TME in clinical and preclinical studies. We further showed that anti-CTLA-4 therapy in combination with iExoKras^G12D^ lends itself to significant anti-tumor response, but not when combined with anti-PD1 therapy. It may reflect a unique mechanism associated with the immune microenvironment remodeling by oncogenic Kras suppression following iExoKras^G12D^ treatment. Notably, inhibition of both Tregs as well as Teffs are critical for anti-CTLA-4 mediated anti-tumor response^25^. Influx of Tregs with iExoKras^G12D^ therapy may open a novel therapeutic window for anti-CTLA-4 therapy in PDAC. iExoKras^G12D^ combination with anti-CTLA-4 therapy leads to emergence of TLS.^26^ Leveraging the information from this clinical trial, the pre-clinical studies offer insights for the design of Phase II iExoKras^G12D^ combination trial with ipilimumab.

## Material and Methods

### Exosomes source and production

GMP iExoKras^G12D^ were derived from the culture supernatant of human bone marrow mesenchymal stromal cells. The procedures to produce GMP-grade iExoKras^G12D^ were detailed previously^17^. Briefly, conditioned media from human bone marrow mesenchymal stromal cells grown in a bioreactor was subjected to filtration and centrifugation. The exosomes were characterized by NanoSight^TM^ NS300 (Malvern) and flow cytometry analyses, and subsequently electroporated with siRNA^17^. MicroBCA analysis was used to define GMP iExoKras^G12D^ doses, expressed as mass of exosomal protein. Flow cytometry analyses of GMP iExoKras^G12D^ was also detailed in prior report, and gating strategy is shown in **Supplementary** Figure 1B.

### Clinical study design and patient population

The iEXPLORE (iExoKras^G12D^ in Pancreatic Cancer, NCT03608631) clinical trial is a first-in-human, open-label, single-center Phase I study of mesenchymal stromal cells-derived exosomes with *KRAS^G12D^* siRNA evaluating the safety, tolerability, and target engagement in patients with metastatic pancreatic ductal adenocarcinoma harboring the KRAS^G12D^ mutation. The study was conducted according to the principles of Good Clinical Practice with prior approvals obtained from the Institutional Review Board (IRB) of MD Anderson’s Human Research Protection Program (HRPP) and with investigational new drug (IND) authorization granted by the Food and Drug Administration (FDA).

The study enrolled patients with pre-defined criteria, including 18 years of age on the day of consenting to the study, and histologically confirmed metastatic PDAC harboring *KRAS^G12D^* mutation, as informed from any previous routine molecular profiling (e.g. Foundation One) of tissue or blood. Patients had documented progression of disease on one or more lines of systemic therapy. If with stable disease, patient must have had completed at least 4 months of chemotherapy with cytotoxic therapy. Patients presented with an Eastern Cooperative Oncology Group (ECOG) performance status of 0 or 1 and adequate organ functions. Women of childbearing potential (WOCBP, defined as not post-menopausal for 12 months or no previous surgical sterilization) had a negative serum pregnancy test within one week prior to initiation of treatment and requested to be using an adequate method of contraception to avoid pregnancy throughout the study and for up to 12 weeks after the last dose of study drug to minimize the risk of pregnancy. Exclusion criteria included concurrent severe and/or uncontrolled medical conditions that could compromise participation in the study, such as unstable angina, myocardial infarction within 6 months, unstable symptomatic arrhythmia, uncontrolled diabetes, serious active or uncontrolled infection, pregnancy or lactation, known CNS disease (except for treated brain metastasis, unless the patient was treated by neurosurgical resection or brain biopsy taken within 3 months prior to Day 1). Patient demographic data are detailed in **Figure 2C** and **Figure 3A**.

The clinical product starting dose of 0.150 mg of exosomal protein (which included an equivalent 0.150 mg of siRNA) was informed by non-toxic dosing in preclinical studies and applying conversion to human equivalent dosing (**Table 1**). iExoKras^G12D^ were generated by the Stem Cell Transplantation and Cellular Therapy Laboratory at MD Anderson according to Good Manufacturing Practice (GMP) and clinical grade siRNA was purchased from City of Hope RNA synthesis core. The iExoKras^G12D^ diluent was PlasmaLyte. Infusion rate of iExoKras^G12D^ was set at 2 mL/min, with infusion time ranging from 5min to 64min for increasing dose levels. Each iExoKras^G12D^ infusion was followed by PlasmaLyte infusion (100 mL over the course of approximately 30min). The initial Phase Ia study enrolled 3 patients at the dose levels (Dose Levels 1, 2, and 3) listed in **Figure 2A**. Given that no toxicity was observed, the Phase Ib study enrolled patients in an accelerated titration design, with a single patient at each of the increasing dose levels (Dose Levels 4, 5, 6). Phase Ib enrollment included capture of pre- and post-treatment tissue biopsies (**Figure 2B**). Toxicity scoring followed the Common Terminology Criteria for Adverse Events (CTCAE) Version 5.0 for toxicity and adverse event reporting. Dose limiting toxicity (DLT) were defined as grade 3 hematologic toxicity with bleeding, grade 4 thrombocytopenia, grade 4 neutropenia of ≥7 days duration or ≥ grade 3 neutropenia of any duration with fever ≥ 38.5°C, grade 2 or greater autoimmune reaction, grade 2 or higher hypersensitivity reaction, grade 4 or higher infusion-related reaction, grade 3 or 4 events, excluding alopecia, nausea, vomiting, and/or diarrhea, unless these occur despite maximal prophylaxis and/or treatment. No DLT were encountered and MTD was not reached.

Clinical response was measured with CT imaging according to RECIST 1.1 criteria, approximately at every 3 cycles, as shown in **Figure 2B**. Target lesions and non-target lesions were ascertained, with Partial Response (PR) defined as at least a 30% decrease in the sum of diameters of target lesions (taking as reference the baseline sum diameters); Progressive Disease (PD) was defined by at least a 20% increase, and Stable Disease (SD) defined as neither sufficient shrinkage to qualify for PR nor sufficient increase to qualify for PD. Appearance of new lesion(s) (NL) also defined PD.

### Non-human primate (NHP) preclinical studies

All procedures were reviewed and approved by the Institutional Animal Care and use Committee (IACUC) at MD Anderson Cancer Center (MDACC) and NHPs were housed in AAALAC-accredited facility at Michale E. Keeling Center for Comparative Medicine and Research (KCCMR) at Bastrop, TX. Two preclinical studies are reported, using GMP iExoKras^G12D^, and prospectively written protocols were followed. Experiment 1 studied the toxicology of GMP iExoKras^G12D^ (**Figure 1E**). Experiment 2 studied the biodistribution of GMP iExoKras^G12D^ (**Figure 1G**).

*Experiment 1*: GMP-iExoKras^G12D^ was administered to adult female and male Rhesus macaques (*Macaca mulatta*) of Indian origin (4.6 to 6.1 kg body weight, 4-5 years old). Rhesus macaques were transferred from the KCCMR Rhesus Macaque Breeding and Research Resource Colony (RMBRR) to study housing acclimation prior to initiation of the pre-clinical study and were randomly assigned to 2 groups. NHP (13-069) in group 1 received PlasmaLyte (diluent, control), and NHPs in group 2 received 0.15 mg GMP iExoKras^G12D^. All doses were administered in 2 ml volume, intravenously at a peripheral venous access point, and flushed with 0.5 ml volume of PlasmaLyte. A total of 9 doses were administered over the course of 42 days (see **Figure 1E**). The NHPs were euthanized one week after the final dose. Body weight (longitudinal and at experimental endpoint), organ weights, chemistry and hematology panels were completed. Gross necropsy findings and histopathological examination of tissues were performed following the proceeding standardized by the KCCMR.

*Experiment 2*: DiR and PKH-67 labeled GMP-iExoKras^G12D^ were administered to adult (approximately 7 years of age) male Rhesus macaques (*Macaca mulatta*) of Indian origin (∼6 kg body weight). For DiR labeling of GMP-iExoKras^G12D^, exosomes were diluted to a final concentration of 5 billion exosomes per 1 ml of PBS and 1 µl of XenoLight DiR (Perkin Elmer, mg/mL) added. The samples were incubated for 15min at 37°C followed by incubation at 4°C for 5min. Labeled exosome samples were then diluted in 10 ml of PBS and centrifuged in a SW41Ti rotor (Beckman Coulter) at 200,000 x g for 3h at 4°C. PKH-67 labeling was performed according to manufacturer’s instructions (Sigma Aldrich). Briefly, exosomes were diluted to a final concentration of 5 billion exosomes per 1 ml in diluent C and 4 µl of PKH-67 added. Exosomes were incubated for 5min at room temperature followed by addition of 2 ml of 1% BSA in water. Samples were then diluted in 8 ml PBS and centrifuged in a SW41Ti rotor at 200,000 x g for 3h at 4°C. Following centrifugation, the supernatant was removed, the exosome pellet resuspended in PlasmaLyte, and exosome concentration evaluated by Nanosight^TM^ NS300 for both DiR and PKH labeled samples.

Rhesus macaques were transferred from the KCCMR RMBRR to study housing acclimation prior to initiation of the pre-clinical study and were randomly assigned to 3 groups. NHP (13-048) in group 1 received GMP iExoKras^G12D^ (siRNA Kras^G12D^) labeled with PKH-67, intravenously, NHP (13-052) in group 2 received GMP iExoKras^G12D^ (siRNA Kras^G12D^) labeled with DiR, intravenously, and NHP (13-072) in group 3 received GMP iExoKras^G12D^ (siRNA Kras^G12D^) labeled with DiR, intraperitoneally. All doses for the biodistribution studies contained approximately 70 mg of exosomal protein (or approximately 70 billion exosomes) administered in 2.5 mL volume (**Table 1**). Rhesus macaques were euthanized 24h post dose with urine, blood, and organs collected. Organs were processed for formalin fixing and paraffin embedding, snap frozen tissues, OCT embedded tissues, and fresh tissue for IVIS imaging. For H&E analysis of paraffin tissues, 5 µm sections were cut and stained for H&E using ST Infinity H&E staining system (Leica) and Leica Autostainer XL according to manufacturer’s instructions.

### NHP tissue analyses

For tissue analyses of DiR^+^ GMP iExoKras^G12D^ biodistribution in NHP, approximately 0.2 to 1 cm^3^ NHP tissue pieces were evaluated for DiR^+^ GMP iExoKras^G12D^ accumulation using In Vivo Imaging System (IVIS Spectrum, Perkin Elmer). Tissues were imaged 24h post injection using a 710 nm excitation filter and 780 nm emission filter with background fluorescence removed by subtracting the background measurements at time of imaging. Tissues were compared to the negative control (NHP 13-048, injected with PKH-67 labeled exosomes).

For microscopic analyses of PKH-67^+^ GMP iExoKras^G12D^ biodistribution in NHP, frozen sections (5 µm) were cut from tissues embedded in optimal cutting temperature (OCT) medium. Sections were stained with 0.25 µg/ml DAPI in TBS for 5min then mounted with coverslips using Fluoroshield (Sigma). Slides were imaged on a Keyence BZ-X710 fluorescent microscope and images stitched together in BZ-X Analysis software (Keyence) to generate a whole tissue image. Threshold values for green fluorescence were established based on negative control slides (NHP 13-052, injected with DiR-labeled exosomes) for each tissue and applied to stitched images in ImageJ. After applying the established threshold values to images, green fluorescence integrated density was quantified within tissue area determined by DAPI positive area. The integrated density of control slides was subtracted to calculate a normalized integrated density for each tissue.

For high resolution imaging of PKH-67 labeled exosomes, DAPI labeled sections (as described above) were imaged with a Zeiss LSM800 confocal microscope equipped with a 63x Plan-Apochromat objective. For the images presented in **Supplementary** Figure 5B, a z-stack spanning the entire tissue thickness at 1 µm intervals was acquired at 20x magnification and the images presented as a maximum intensity projection of the entire z-stack.

### Murine preclinical studies

All procedures were reviewed and approved by the Institutional Animal Care and use Committee (IACUC) at MD Anderson Cancer Center (MDACC) and mice were housed in AAALAC-accredited facility at MDACC. All mice received LabDiet 5053 *ad libitum* and were housed at 21-23°C, 40-60% humidity and 12h light/dark cycle.

Two preclinical studies were reported. The toxicology study (**Figure 1A**) employed GMP iExoKras^G12D^ administered intravenously to adult wild-type C57BL/6 albino females (age 25-30 weeks). Mice were purchased from Jackson Laboratories and allowed to acclimate for 2 weeks or more prior to initiation of the preclinical study. Ten mice were randomly assigned to 4 groups each. Mice in group 1 received PlasmaLyte (diluent, control), and mice in group 2 to 4 received increasing dose of GMP iExoKras^G12D^. Group 2 was administered 0.66 µg of iExoKras^G12D^ per dose, Group 3 was administered 1.32 µg of iExoKras^G12D^ per dose, and Group 4 was administered 2.65 µg of iExoKras^G12D^ per dose. All doses were administered in 50 µl volume, intravenously via the retro-orbital plexus. A total of 9 doses were administered over the course of 6 weeks (see **Figure 1C**). The mice were euthanized within one week after the final dose. Body weight (longitudinal and at experimental endpoint), organ weights, chemistry and hematology panels were completed. Gross necropsy findings and histopathological examination of tissues were performed following the proceeding standardized by the Department of Veterinary Medicine and Surgery, Division of Veterinary and Comparative Pathology at MDACC.

*P48-Cre, LSL-Kras^G12D/+^, Tgfbr2^F/F^* (KTC) GEM tumor tissue sections used for analysis in **Figure 6A-C** were obtained from a previously reported study^16^. For studies with T cell depletion mice, we crossed KPC (*Pdx1-Cre, LSL-Kras^G12D/+^, Trp53^R172H/+^*)^27^ mice with CD4^−/−^ (*Cd4^tm1Mak^*)^28^ depleted of CD4^+^ T cells or CD8^+^ T cells CD8^−/−^ (*Cd8^tm1Mak^*)^29^. The combination preclinical study using GMP iExoKras^G12D^ and ICB antibodies utilized orthotopic PKC-Hy19636 (derived *P48Cre, LSL-Kras^G12D/+^, Trp53^F/+^* mice)^30^ cells that were injected in the tail of the pancreas of C57BL6/J mice (0.5 x10^6^ cells in 20 ml PBS using 27-gauge Hamilton syringe, **Figure 6D**). Both male and female adult mice were used and purchased from Jackson laboratories. For checkpoint blockade immunotherapy experiments, anti-CTLA-4 (BioXcell, Clone 9H10) or anti-PD-1 (BioXcell, Clone 29F1.A12) were injected three times per week (first dose 200 mg, followed by 2 doses of 100 mg intraperitoneal injections) in a final volume of 100 ml diluted in PBS. The control mice received a cocktail of isotype antibodies (Rat IgG2a (BioXcell, Clone 2A3) and Syrian hamster IgG (BioXcell, BE0087)) and control exosomes (non-electroporated with siRNA) in the same dosage, route and frequency as the treatment groups. For iExoKras^G12D^ treatments, 10^9^ GMP iExoKras^G12D^ (equivalent to approximately 1 µg) were injected everyday intraperitoneally in the orthotopic PKC mice and every other day in the KTC GEM study. For assessments of tumor volumes, mice were imaged at indicated time points using Bruker 7T MRI at the MD Anderson Small Animal Imaging Facility (SAIF). The mice were sacrificed at indicated time points for age-matched analysis.

### Immunolabeling studies

For samples from the clinical trial, serial sections were cut through the block to collect 5 μm formalin fixed paraffin embedded (FFPE) sections. H&E analysis was performed on slides every 100 μm to select serial sections with optimal tumor tissue for immunostaining analysis. Immunohistochemistry for phosphorylated ERK (pERK, **Figure 4F**) were carried on 5 μm thick FFPE sections using an automated protocol (Leica Bond RX; Leica Biosystems Nussloch GmbH). The tissue sections were deparaffinized and rehydrated following the Leica Bond protocol. Antigen retrieval was performed for 20min with Bond Solution #1 (Leica Biosystems, equivalent to citrate buffer, pH 6.0). The primary antibody [Phospho-p44/42 MAPK (Erk1/2) (Thr202/Tyr204) (D13.14.4E) XP® Rabbit monoclonal antibody (Cell signaling, Cat#4370), dilution 1:400, Cell signaling, Cat#4370] was incubated for 15min at room temperature. The primary antibody was detected using the Bond Polymer Refine Detection kit (Leica Biosystems) with DAB as chromogen. The slides were counterstained with hematoxylin, dehydrated, and coverslipped.

Immunofluorescence analysis was performed on 5 μm thick FFPE sections using Tyramide signal amplification (TSA) after deparaffinization and hydration, following which antigen retrieval was performed for 20min at 98°C in Tris-EDTA buffer (pH-8.6) in a steamer. Next, the sections were incubated in 3% H_2_O_2_ for 10min at room temperature, 1.5% bovine serum albumin (BSA) for 1h and primary antibody at indicated concentrations for 3h (**Supplementary Table 1**). Following addition of the indicated secondary polymer (30min at RT), opal reagents were used at a 1:100 concentration (30min at RT) for TSA staining. Subsequently, antigen retrieval was performed before use of the next primary antibody. The slides were mounted using Fluoroshield^TM^ with DAPI (Sigma Aldrich F6057). For single stained IHCs, DAB reagent (5 to 15min) was added following incubation with the secondary polymer, counterstained with hematoxylin, dehydrated and mounted for analysis. For quantification of immune infiltrates, representative images were taken from 3-5 high power fields (20x magnification) per biological replicate to count the number of immune cells and average of multiple images is plotted as a replicate. We used Zeiss Axio Imager Z.2 for IF imaging. For quantification of CK19 and pERK immunostainings, average pixels positivity of 5 high power field images were used for quantification using Leica microsystems DFC295 microscope for brightfield imaging.

### Digital PCR analyses for circulating Kras^G12D^ DNA

We previously reported on the anti-tumor efficacy of GMP iExoKras^G12D^ with and without gemcitabine in an orthotopic model of pancreatic cancer^17^. The blood of mice from the previously published experiment (Reference^17^, *Figure 6E & Supplementary* Figure 8C) was analyzed for the detection of Kras^G12D^ mutation in circulating DNA. Briefly, blood samples were collected from tumor bearing mice (KPC689 orthotopic tumors with Kras^G12D^ mutation) one day prior to the start of treatment (pre-treatment/pre-tx) and 9 days after treatment was started (post-treatment/post-tx) (see **Figure 4A**). Murine blood samples were collected and left at room temperature for 4h followed by centrifugation at 3,000 x g for 5min to collect serum. cfDNA from the serum for dPCR were extracted by using QIAamp DNA Micro Kit and DNA concentration was measured by Qubit dsDNA HS kit according to manufacturer’s instructions. The probe assay ID used was AH6R5PI (KRAS G12D c35G>A (WT:C —> MUT:T). 6.5 µl of DNA was added regardless of DNA concentration.

Approximately 1.5 to 4 ml plasma samples for patients on the clinical trial were also analyzed. ctDNA isolated from 0.5 ml of patient plasma using the cfPure V2 Cell Free DNA Extraction Kit (Biochain) according to manufacturer’s instructions. TaqMan Liquid Biopsy dPCR Assay (Invitrogen A44177, Assay ID Hs000000051_rm) in conjunction with Quant Studio 3D dPCR 20K was used to amplify 1 or 2 ng of ctDNA and quantify (averaging values for 1 and 2 ng) *KRAS^G12D^* and *KRAS* (wildtype/unmutated allele) in ctDNA.

The analysis of the digital PCR data was performed with the manufacturer’s software (QuantStudio 3D Analysis Suite). The setting of detection limit used was the same as previously reported^31^. The results were expressed as a percent of the ratio of Target/Total (Kras^G12D^/(Kras^G12D^+Kras^WT^)).

### Quantitative RT-PCR

For *in vitro* experiments to determine Kras^G12D^ knockdown efficacy of iExoKras^G12D^, 10^5^ Panc-1 or KPC-689 cells were seeded in 6 well plates (ThermoFisher, 140685) in 10% FBS DMEM with PS in a final volume of 2 ml per well overnight. Subsequently, 10^9^ iExoKras^G12D^ were added in serum free DMEM in a final volume of 1 ml and RNA was extracted from the cell lines using PureLink^TM^ RNA Mini Kit (Invitrogen, 12183025) as per the manufacturer’s instructions at indicated time points for downstream analysis. RNA concentrations were determined using spectrophotometer (Nanodrop 2000) and 500 μg to1 mg RNA was used for reverse transcription using High-Capacity cDNA Reverse Transcription Kit (ThermoFisher Scientific, 4368814). The cDNA was further diluted 1:20 for qPCR analysis with SYBR^TM^ green master mix (Applied Biosystems^TM^, A25742). Primers used for qPCRs include:

*_KrasG12D_*

F: 5’-ACTTGTGGTGGTTGGAGCAGA-3’ R: 5’-TAGGGTCATACTCATCCACAA-3’

*Fas:*

F: 5’-TATCAAGGAGGCCCATTTTGC-3’ R:5’-TGTTTCCACTTCTAAACCATGCT-3’ *Il15*:

F: 5’-ACATCCATCTCGTGCTACTTGT-3’ R: 5’-GCCTCTGTTTTAGGGAGACCT-3’ *18s*:

F: 5’-GTAACCCGTTGAACCCCATT-3’

R: 5’-CCATCCAATCGGTAGTAGCG-3’

*Gapdh:*

F: 5’-AGGTCGGTGTGAACGGATTT-3’

R: 5’-TGTAGACCATGTAGTTGAGGTCA-3’

Gene expression for listed genes was normalized to *18s* or *Gapdh* and fold change was determined by ratios of 2^-dCt^ values. Statistical comparisons were performed on dCt values.

### Chromatin immunoprecipitation

For ChIP experiments, 0.5 to 1 x 10^6^ cells per immunoprecipitation (IP) condition from iExo and ScrExo [2 doses (10^9^ exosomes/dosage) administered 24h apart] of KPC-689 cells were collected on day 3 by trypsinization. Cross-linking was performed by shaking cells in a suspension of 1% methanol free formaldehyde (ThermoFischer, Cat# 28908) in complete medium at room temperature for 10 to 12min. Subsequently, the solution was mixed with 125 mM glycine and incubated at room temperature for 5min. The cells were washed twice with ice cold PBS (with 1 mM PMSF) and the resulting cell pellets were stored at –80^°^C for downstream analysis. ChIP assays were performed at the MD Anderson’s Epigenomics Profiling Core with some modifications to previously described high-throughput ChIP protocol^32^. For histone ChIPs, H3K27ac, Abcam – Cat# ab4729; H3K27me3, Diagenode – Cat# C15410195 and H3, Abcam – Cat# ab1791 were used. For ChIPs for non-histone antibodies (DNMT1, Novus Biologicals, Cat# NB100-56519 and EZH2, Cell Signaling Technology, Cat# 5246S) were used. Subsequently, qPCR assays were performed using primers for the promoter/transcription start site using the respective primers (FAS TSS (Histone ChIP)-F: 5’-CTGCCTCTGGTAA GCTTTGG-3’ R:5’-CAGCCACATCTGGAATCTCA-3’; FAS promoter site (DNMT1, EZH2 ChIP)-F: 5’-CCC TGTATTCCCATTCATCG-3’ and R: 5’-ACTAGGGG AGGGGACAGAAA-3’). Normalized transcript levels (to IgG or histone H3) were used for statistical comparisons.

### Electron microscopy

<u>Scanning electron microscopy</u> – GMP iExoKras^G12D^ was submitted to the High Resolution Electron Microscopy Facility for processing and imaging. Fixed samples were washed in 0.1 M sodium cacodylate buffer and treated with 0.1% Millipore-filtered cacodylate buffered tannic acid, postfixed with 1% buffered osmium, and stained *en bloc* with 1% Millipore-filtered uranyl acetate. The samples were dehydrated in increasing concentrations of ethanol, infiltrated, and embedded in LX-112 medium. The samples were polymerized in a 60°C oven for approximately 3 days. Ultrathin sections were cut in a Leica Ultracut microtome (Leica, Deerfield, IL), stained with uranyl acetate and lead citrate in a Leica EM Stainer, and examined in a JEM 1010 transmission electron microscope (JEOL, USA, Inc., Peabody, MA) at an accelerating voltage of 80 kV. Digital images were obtained using AMT Imaging System (Advanced Microscopy Techniques Corp, Danvers, MA). All pictures were captured with the help of the facility.

<u>Cryo-electron microscopy</u> – GMP iExoKras^G12D^ was submitted to the CryoEM Core at Baylor College of Medicine for processing and imaging. The surfaces of QUANTIFOIL holey carbon grids were plasma cleaned for 20 seconds inside an Ernest Fullam Glow Discharge. The grids were transferred to a VitroBot Mark IV plunge freezer where 2.5 µl of the iExoKras^G12D^ sample and 1 µl of AURION 10 nm gold tracers were applied to the carbon-side of the grid before immediately being plunged into liquid nitrogen. The vitrified sample was transferred to a JEOL 2200FS operating at 200 kV equipped with a Direct Electron DE-64 detector. Micrographs were collected at 8k x 8k resolution with a final A/pixel of 2.18. The approximate total electron dose per micrograph was 20 e-/A^2. The micrograph movies were motion corrected and gain-normalized using EMAN2^33^.

**Supplementary Figure Legend:**

**Supplementary Figure 1. GMP iExoKras^G12D^ characterization**

**A.** GMP iExoKras^G12D^ particle size and concentration by NanoSight^TM^. **B**. Gating strategy for GM iExoKras^G12D^ bound to beads. **C**. Histogram of flow cytometry analysis of GMP iExoKras^G12D^ for the indicated markers. **D**. Representative SEM of GMP iExoKras^G12D^, scale bars listed. **E**. Representative cryoEM of GMP iExoKras^G12D^. Scale bar: 100 nm or otherwise listed.

**Supplementary Figure 2. Safety profile of systemic GMP iExoKras^G12D^ in mice**

**A-B**. Percent change in body weight (A) and listed organs (B) for the listed groups of mice, PT: pretreatment. 1X, 2X, and 4X exosomes reflect dosing corresponding to clinical trial dosing, see also **Table 1**, n=10 mice per group. **C-D**. Chemistry and hematology tabulated results (C) or graphical representation (D) for the listed groups. Data presented as scatter plot about the mean. One-way ANOVA with Tukey’s multiple comparisons test (A-B) or Dunnett’s multiple comparisons test (C-D). *P<0.05; **P<0.01; ns, not significant.

**Supplementary** Figure 3**. Safety profile of systemic GMP iExoKras^G12D^ in Rhesus macaques A-B**. Normalized body weight over time (A) and organ/body weight (BW) ratio of the listed organs for each Rhesus macaque. **C**. Graphical representation of hematology and chemistry analysis of the listed Rhesus macaque.

**Supplementary** Figure 4**. Labeled exosomes biodistribution in Rhesus macaque** Representative images of pancreas, liver, and brain tissue section from Rhesus macaque with nuclear staining (DAPI) and PKH labeled exosomes with digital zoom inset.

**Supplementary Figure 5. High magnification images of labeled exosomes biodistribution in Rhesus macaque**

**A.** Representative high magnification images of the listed organs in Rhesus macaque with nuclear staining (DAPI) and PKH labeled exosomes. The panel displays negative control (unlabeled exosomes) next to PKH^+^ label exosomes-treated Rhesus macaque; this panel is also displayed in **Figure 1J**. **B**. Representative confocal microscopy images of the listed organs from Rhesus macaque.

Supplementary Figure 6. Biodistribution of exosomes in Rhesus macaque

**A.** Representative H&E stained images of the listed organs of each Rhesus macaque. **B**. Normalized relative luminescence unit (RLU) in the listed organs for the detection of DiR^+^ label following i.v. or i.p. DiR^+^ iExoKras^G12D^ treatment (relative to control organ without exposure to DiR^+^ iExoKras^G12D^). 1: unreliability of data due to high RLU in control organ; 2: unreliability of data due to lack of background RLU detection in control organ.

**Supplementary Figure 7. Circulating siRNA capture by dPCR in Rhesus macaque**

**A-B**. Histogram representation (A) and dPCR scatter plot (B) of the percent circulating siRNA dPCR capture relative to total RNA in Rhesus macaque 24h following i.v. or i.p. administration of iExoKras^G12D^. siRNA concentration curve for positive control and range of detection used 27 ng, 270 pg, 27 pg of Kras^G12D^ siRNA and 270 pg of scrambled (scrbl) control siRNA. Rhesus macaque ID listed, refers to **Figure 1G**.

**Supplementary Figure 8. Immunostaining of tissue biopsies**

**A.** Representative images of immunolabeling for CD4, Foxp3, nuclear stain (DAPI) and respective quantification of percentage of CD4^+^Foxp3^+^ and CD4^+^Foxp3^-^ T cells in paired biopsies. Scale bar:100 μm. **B**. Relative *KRAS^G12D^* expression in Panc-1 cells treated with iExoKras^G12D^ stored for 4 years at –80°C. Data are presented as mean (A) and mean +/– SEM (B). Unpaired t test without Welch’s correction, **P<0.01.

**Supplementary Figure 9. Contribution of CD4^+^ and CD8^+^ T cells in iExoKras^G12D^ therapy response in mice**

**A.** Schematic of experimental timeline using orthotopic KPC-689 mice. **B.** Representative images and respective quantification of CD4^+^ T cells (CD4), and CD8^+^ T cells (CD8) by immunolabeling in control and iExoKras^G12D^ treated orthotopic KPC-689 mice, n=4 mice per group. Scale bar: 100 μm. **C**. Schematic of experimental timeline using KPC, KPC CD4^−/−^ and KPC CD8^−/−^ GEMs treated with iExoKras^G12D^. **D**. Representative MRI images and quantification of percent change in tumor burden at indicated time points of KPC, KPC CD4^−/−^ and KPC CD8^−/−^ treated with iExoKras^G12D^, n=5 to 8 mice per group. **E**. Kaplan-Meier survival curves showing disease specific survival of KPC-Control (n=15 mice), KPC-iExo (n=7 mice), KPC CD4^−/−^ control (n=9 mice), KPC CD4^−/−^ iExo (n=7 mice), KPC CD8^−/−^ control (n=10 mice), KPC CD8^−/−^ iExo (n=7 mice). **F** qPCR analysis of *Kras^G12D^*, *Fas* and *Il15* expression following treatment with iExoKras^G12D^ or scrambled (ScrExo, control) (2 doses, 24h apart). RNA extracted at 72h (n=3 wells per group). **G** ChIP assays demonstrating the relative binding of DNMT1, EZH2, H3K27me3, and H3K27ac on FAS promoter in KPC-689 cells in the indicated treatment groups (2 doses, 24h apart). n=4-5 biological replicates per group. Data are presented as mean +/– SD. Unpaired t test without Welch’s correction (B, F, G), one-way ANOVA with Tukey’s/Dunnett’s multiple comparisons test (D), log-rank test (E). *P<0.05; **P<0.01; ***P<0.001; ****P<0.0001; ns, not significant.

**Supplementary Figure 10. TLS in iExoKras^G12D^ and ICB combination treatment**

**A.** Representative H&E (A) and quantification (B) of TLS in the indicated groups. Scale bar: 100 μm. Data are presented as mean +/– SD. Kruskal-Wallis with Dunn’s multiple comparisons test. **P<0.01.

## Author contribution

VSL, MLK, KMM, SY, HS, LMB, ASM and MG contributed to the murine preclinical studies, and VSL, MLK, KMM, SY, HS, RF also contributed to the NHP preclinical studies. AF and SL contributed to data acquisition and analysis. VSL, KKM, ASM, LMSS, CH contributed to human sample analyses. VSL, BGS, JJKL, GV, RTS, AM, MM, EJ, and SP contributed to the clinical trial design and execution. VSL, KKM, RK, BS, SP, AM, ES conceptually designed the strategy for this study, provided intellectual input, and contributed to the writing of the manuscript.

## Data Availability

All data produced in the present study are available upon reasonable request to the authors

## Acknowledgements

The work was partially supported by the Cancer Prevention and Research Institute of Texas. The iExoKras^G12D^ preclinical studies and Phase I trial were supported by the MD Anderson Moon Shot Program and in part by the Translational Molecular Pathology-Immunoprofiling Lab (TMP-IL) Moon Shots Platform. A.M. is supported by the Khalifa bin Zayed Foundation. The work was done using the High-Resolution Electron Microscopy and the Small Animal Imaging Facility Core Facilities at MDACC. The CryoEM data were collected at the Baylor College of Medicine CryoEM ATC, subsidized by CPRIT Core Facility Award RP190602 which also supported acquisition of CryoEM equipment used in this study. R35GM151999 and R01GM080139 to SL in support of CryoEM data collection and analysis. We thank Kenneth Dunner Jr. for help with the electron microscopy analyses; Xunian Zhou for flow cytometry analyses; Swati Gite, Leticia Campos Clemente, Mei Jian, Wei Lu and Khaja Khan for coordination of tissue sample and analysis of chromogenic IHC data; Charles Kingsley, Jorge Delacerda and Houra Taghavi for assistance with MRI imaging. The authors acknowledge the contributions of Dr. Pamela Papadopoulos in MD Anderson Moon Shots Operations for Program Management support and Dr. Mark Hurd in the Ahmed Center for biospecimen collection support.

