## Supplementary Figures for "KRAS^G12D^-Specific Targeting with Engineered Exosomes Reprograms the Immune Microenvironment to Enable Efficacy of Immune Checkpoint Therapy in PDAC Patients"

### Supplementary Figure 1

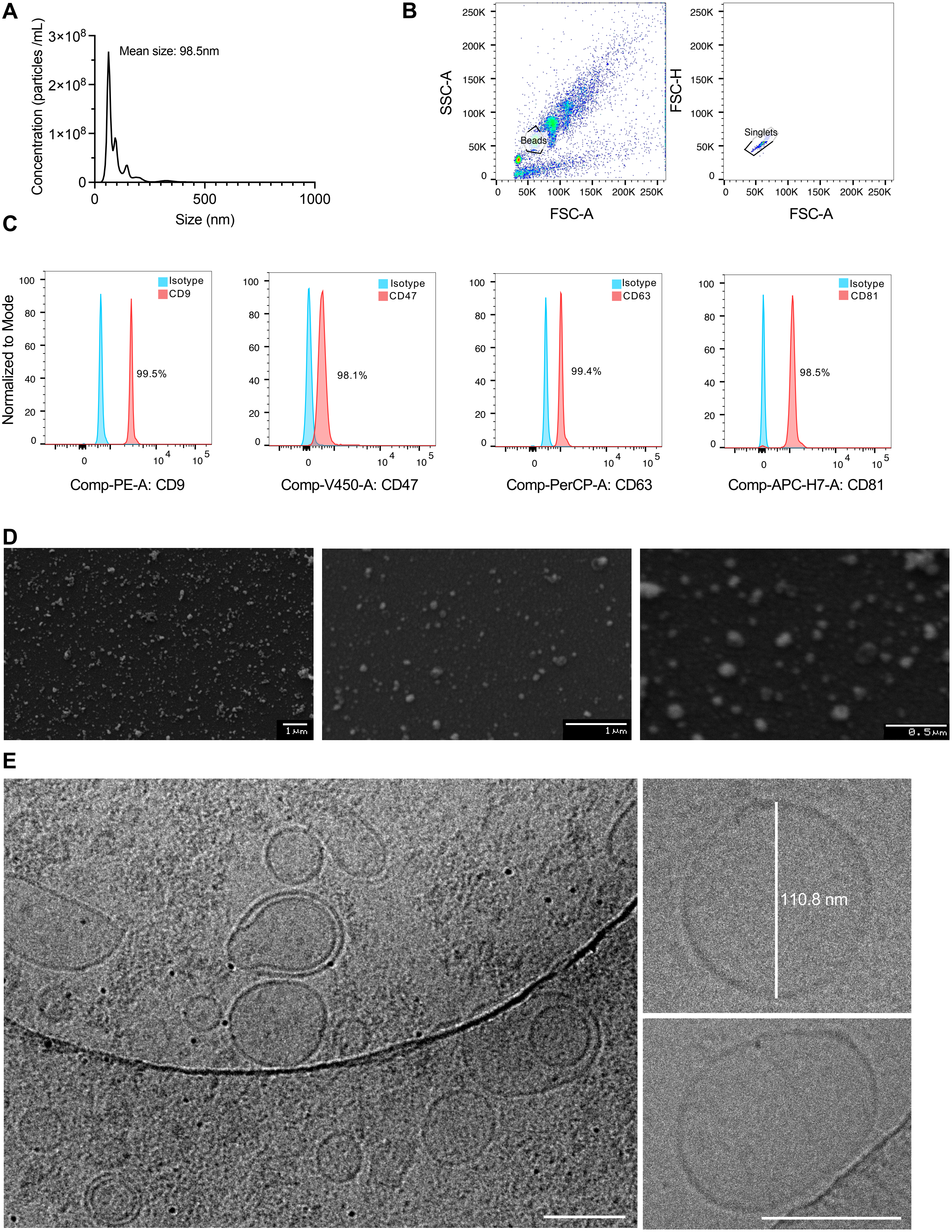

### Supplementary Figure 2

**A**

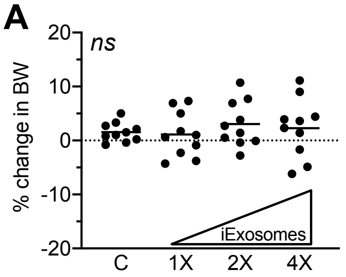


**B**

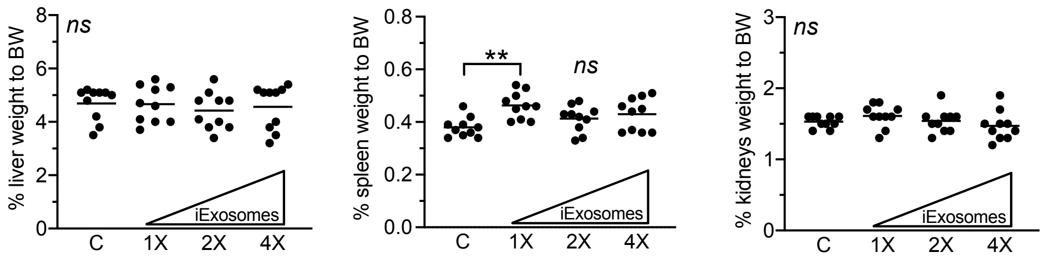


**C**

| Chemistry & Hematology | Control (C) | | iExosomes, 1X | | iExosomes, 2X | | iExosomes, 4X | | Significance | Normal Range (Y/N) |
|---|---|---|---|---|---|---|---|---|---|---|
| | Mean | SD | Mean | SD | Mean | SD | Mean | SD | | |
| Albumin (g/dL) | 4.14 | 0.22 | 3.96 | 0.20 | 3.95 | 0.16 | 3.95 | 0.24 | ns | Y |
| ALP (U/L) | 120.10 | 15.47 | 105.10 | 12.99 | 107.70 | 22.06 | 94.00 | 26.08 | * C vs 4X | Y |
| ALT (U/L) | 191.60 | 227.65 | 155.30 | 213.46 | 157.00 | 128.48 | 130.70 | 132.71 | ns | Y |
| AST (U/L) | 299.20 | 246.55 | 285.00 | 289.87 | 333.80 | 307.52 | 250.40 | 155.74 | ns | Y |
| Total bilirubin (mg/dL) † | <0.2 | n/a | <0.2 | n/a | <0.8 | n/a | <0.3 | n/a | n/a | Y |
| Blood Urea (mg/dL) | 19.13 | 3.65 | 19.26 | 3.21 | 19.61 | 3.75 | 19.13 | 2.62 | ns | Y |
| Calcium (mg/dL) | 10.88 | 0.46 | 10.98 | 0.23 | 10.72 | 0.40 | 11.12 | 0.28 | ns | Y |
| Chloride (mEq/L) | 114.24 | 1.59 | 115.52 | 1.29 | 114.81 | 1.42 | 114.83 | 3.05 | ns | Y |
| Creatinine (mg/dL) † | <0.25 | n/a | <0.24 | n/a | <0.22 | n/a | <0.25 | n/a | n/a | Y |
| Globulin (g/dL) | 1.53 | 0.19 | 1.54 | 0.17 | 1.61 | 0.23 | 1.63 | 0.19 | ns | Y |
| Potassium (mEq/L) | 9.62 | 0.89 | 9.74 | 0.96 | 10.05 | 1.31 | 9.78 | 0.76 | ns | Y |
| LDH (U/L) | 559.89 | 521.23 | 515.10 | 478.23 | 622.70 | 459.19 | 488.89 | 258.31 | ns | Y |
| Sodium (mEq/L) | 153.28 | 1.65 | 155.06 | 2.01 | 156.78 | 7.18 | 154.36 | 1.78 | ns | Y |
| Phosphorus (mg/dL) | 11.46 | 1.51 | 11.05 | 1.09 | 10.25 | 0.82 | 11.06 | 0.79 | ns | Y |
| Glucose (mg/dL) | 311.78 | 84.09 | 271.90 | 52.20 | 275.50 | 41.99 | 310.00 | 68.13 | ns | Y |
| Total protein (g/dL) | 5.68 | 0.33 | 5.50 | 0.29 | 5.57 | 0.28 | 5.59 | 0.25 | ns | Y |
| WBC count (x 10.e3/μL) | 3.82 | 0.98 | 3.92 | 1.42 | 4.15 | 1.13 | 5.18 | 2.36 | ns | Y |
| RBC count (x 10.e6/μL) | 10.36 | 0.57 | 9.99 | 0.53 | 10.23 | 0.45 | 10.04 | 0.48 | ns | Y |
| Hemoglobin (g/dL) | 15.45 | 0.82 | 15.39 | 0.56 | 15.47 | 0.51 | 15.20 | 0.47 | ns | Y |
| Hematocrit (%) | 51.98 | 3.17 | 50.87 | 2.05 | 51.45 | 1.76 | 50.53 | 1.61 | ns | Y |
| MCV (fl) | 50.16 | 1.33 | 50.97 | 1.53 | 50.33 | 1.53 | 50.39 | 1.07 | ns | Y |
| MCH (pg) | 14.92 | 0.37 | 15.41 | 0.49 | 15.14 | 0.55 | 15.14 | 0.37 | ns | Y |
| MCHC (g/dL) | 29.76 | 0.46 | 30.23 | 0.42 | 30.07 | 0.26 | 30.08 | 0.49 | * C vs 1X | Y |
| RDW (%) | 12.95 | 0.28 | 13.26 | 0.80 | 13.20 | 0.55 | 13.34 | 0.58 | ns | Y |
| Platelet count (x 10.e3/μL) | 1175.50 | 145.83 | 976.67 | 221.33 | 1054.80 | 175.71 | 1055.50 | 132.87 | * C vs 1X | Y |
| MPV (fl) | 4.76 | 0.18 | 4.82 | 0.11 | 4.71 | 0.13 | 4.82 | 0.08 | ns | Y |
| Segs (%) | 9.94 | 2.57 | 12.59 | 2.97 | 13.93 | 3.30 | 13.18 | 6.47 | ns | Y |
| Lymphs (%) | 84.21 | 2.62 | 79.70 | 4.08 | 80.43 | 3.54 | 79.98 | 5.90 | ns | Y |
| Monos (%) | 1.42 | 0.57 | 2.22 | 1.33 | 1.70 | 0.56 | 1.69 | 0.55 | ns | Y |
| Eos (%) | 0.64 | 0.26 | 1.30 | 0.97 | 0.95 | 0.25 | 1.49 | 0.49 | ** C vs 4X | Y |
| Basos (%) | 0.76 | 0.31 | 0.98 | 0.47 | 0.81 | 0.41 | 0.92 | 0.53 | ns | Y |
| LUC (%) | 2.53 | 0.82 | 3.13 | 1.21 | 2.18 | 0.99 | 2.76 | 1.20 | ns | Y |
| Segs (x 10.e3/μL) | 0.39 | 0.15 | 0.50 | 0.22 | 0.58 | 0.19 | 0.73 | 0.56 | ns | Y |
| Lymphs (x 10.e3/μL) | 3.21 | 0.83 | 3.14 | 1.20 | 3.34 | 0.95 | 4.09 | 1.84 | ns | Y |
| Monos (x 10.e3/μL) | 0.05 | 0.03 | 0.09 | 0.05 | 0.07 | 0.03 | 0.09 | 0.05 | ns | Y |
| Eos (x 10.e3/μL) | 0.02 | 0.01 | 0.05 | 0.02 | 0.04 | 0.02 | 0.08 | 0.06 | ** C vs 4X | Y |
| Basos (x 10.e3/μL) | 0.03 | 0.01 | 0.04 | 0.01 | 0.03 | 0.01 | 0.04 | 0.02 | * C vs 4X | Y |
| LUC (x 10.e3/μL) | 0.09 | 0.03 | 0.11 | 0.05 | 0.09 | 0.05 | 0.16 | 0.12 | ns | Y |

**D**

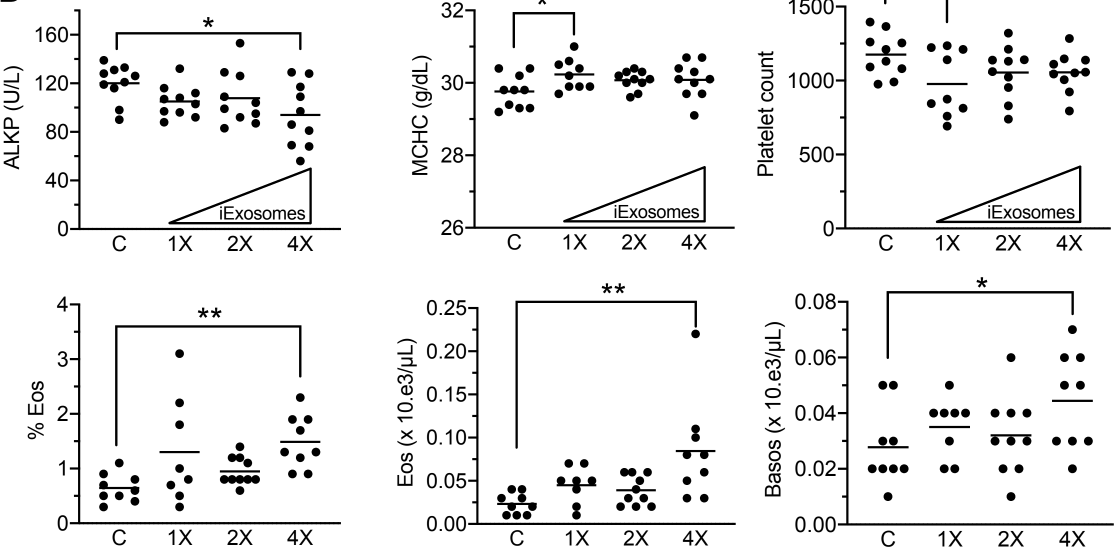

### Supplementary Figure 3

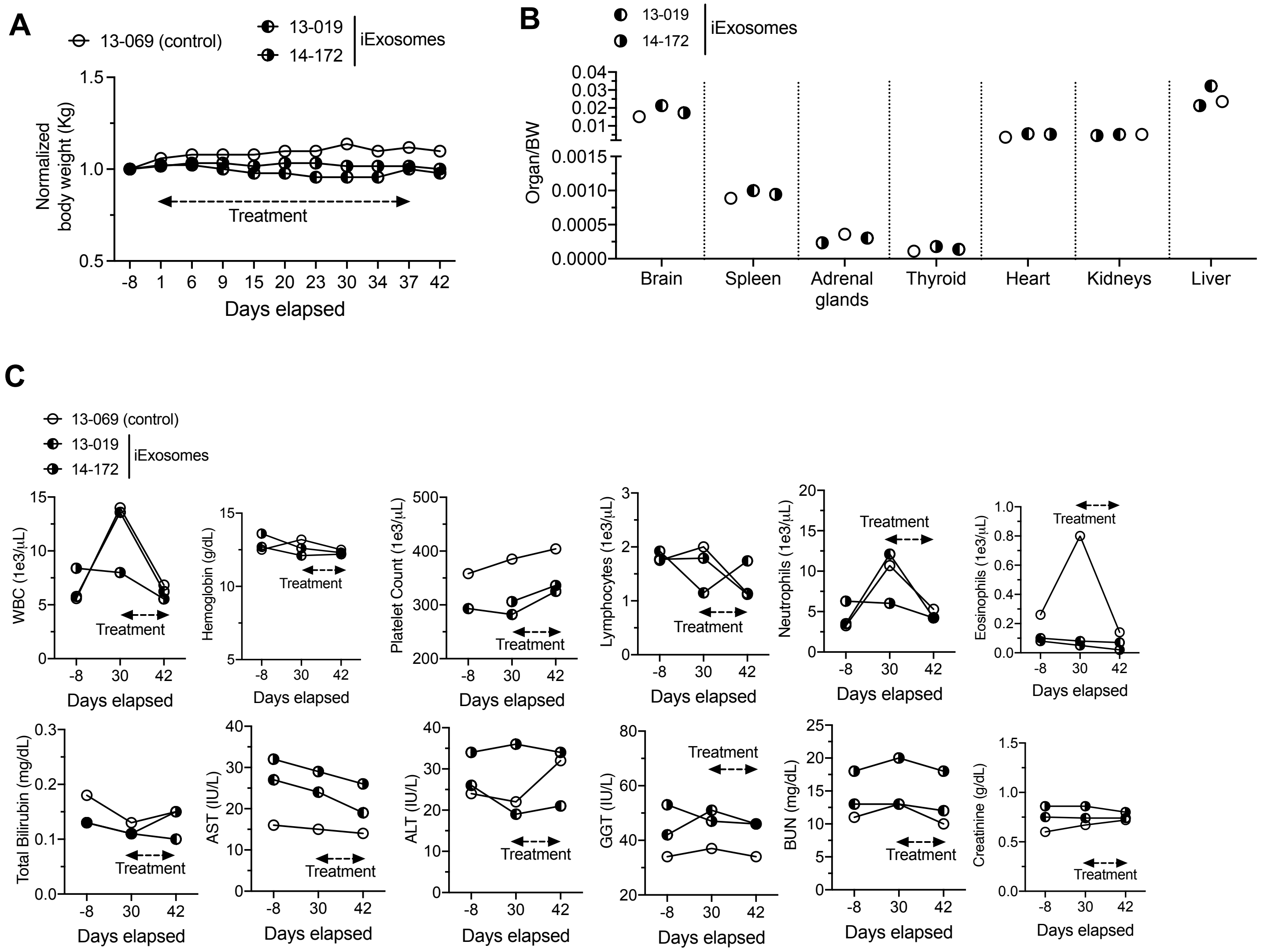

### Supplementary Figure 4

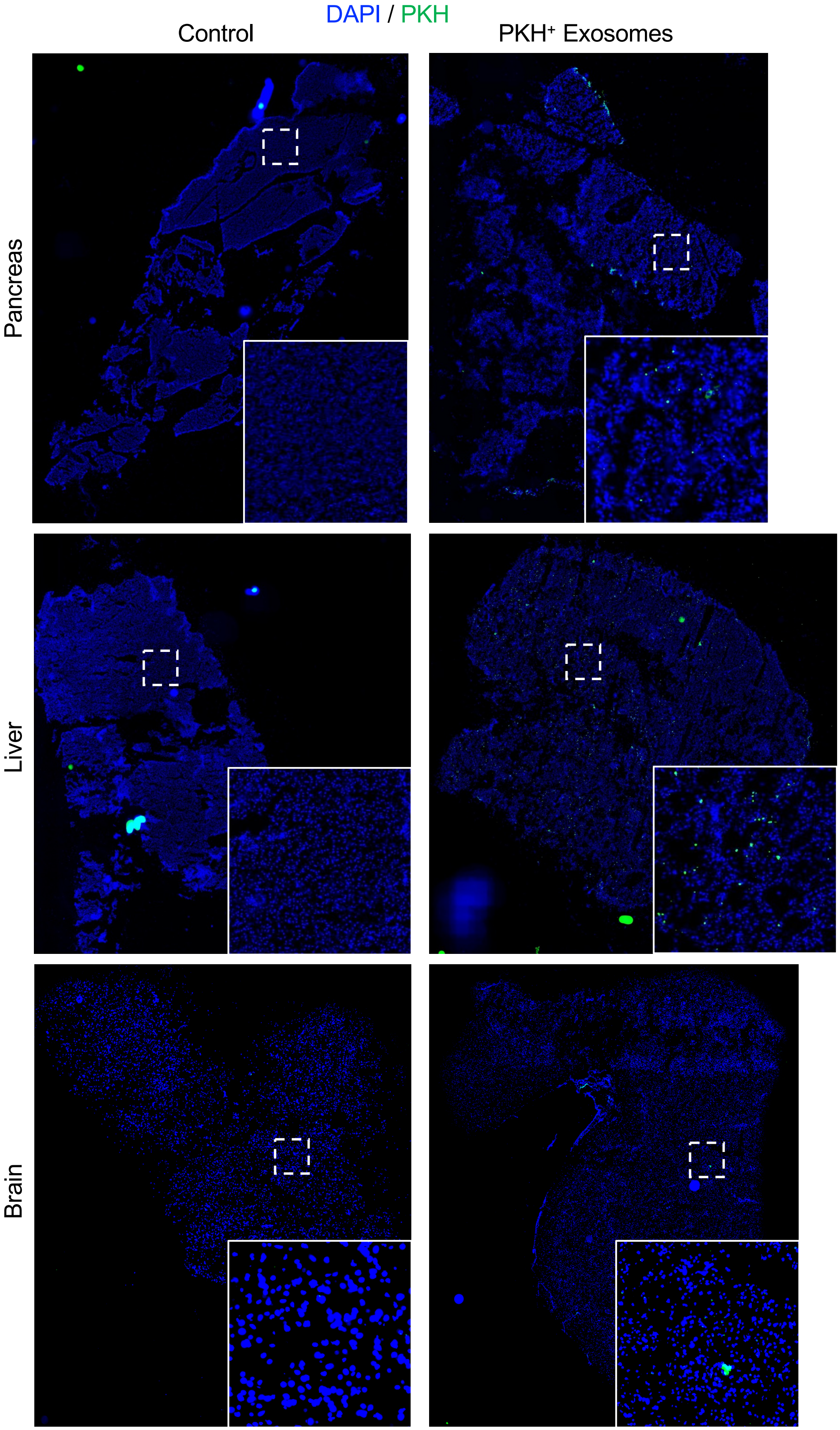

### Supplementary Figure 5

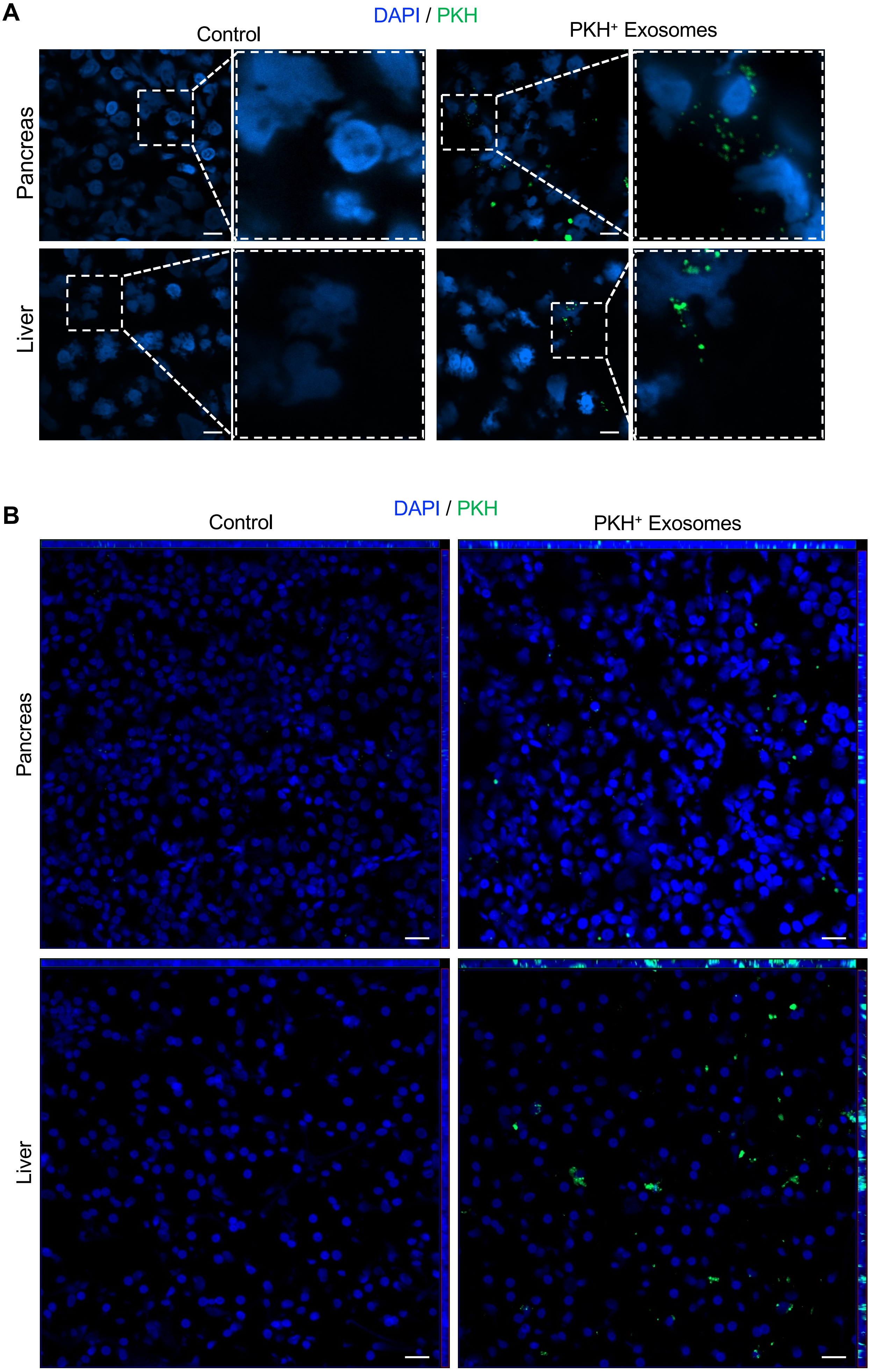

#### Supplementary Figure 6

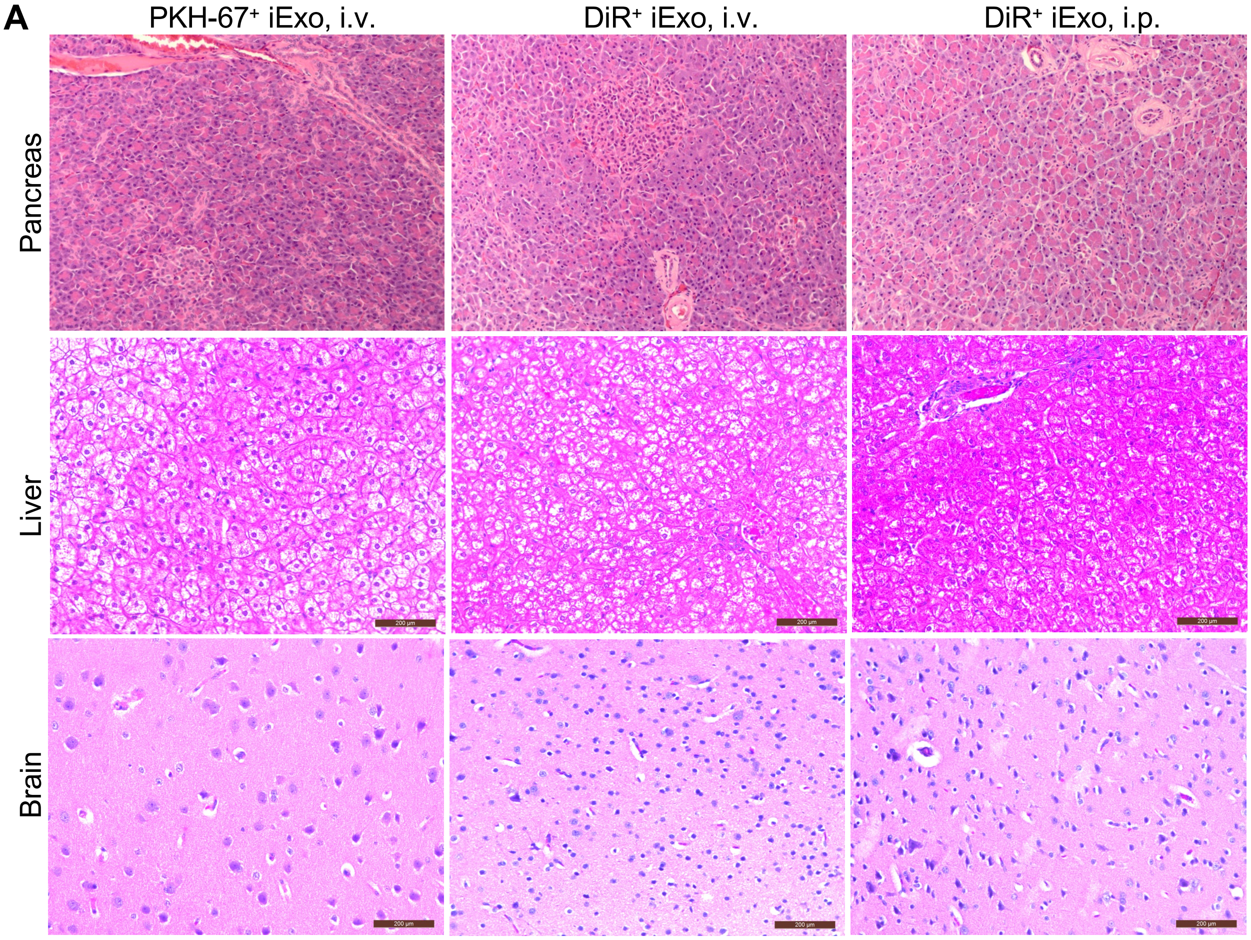


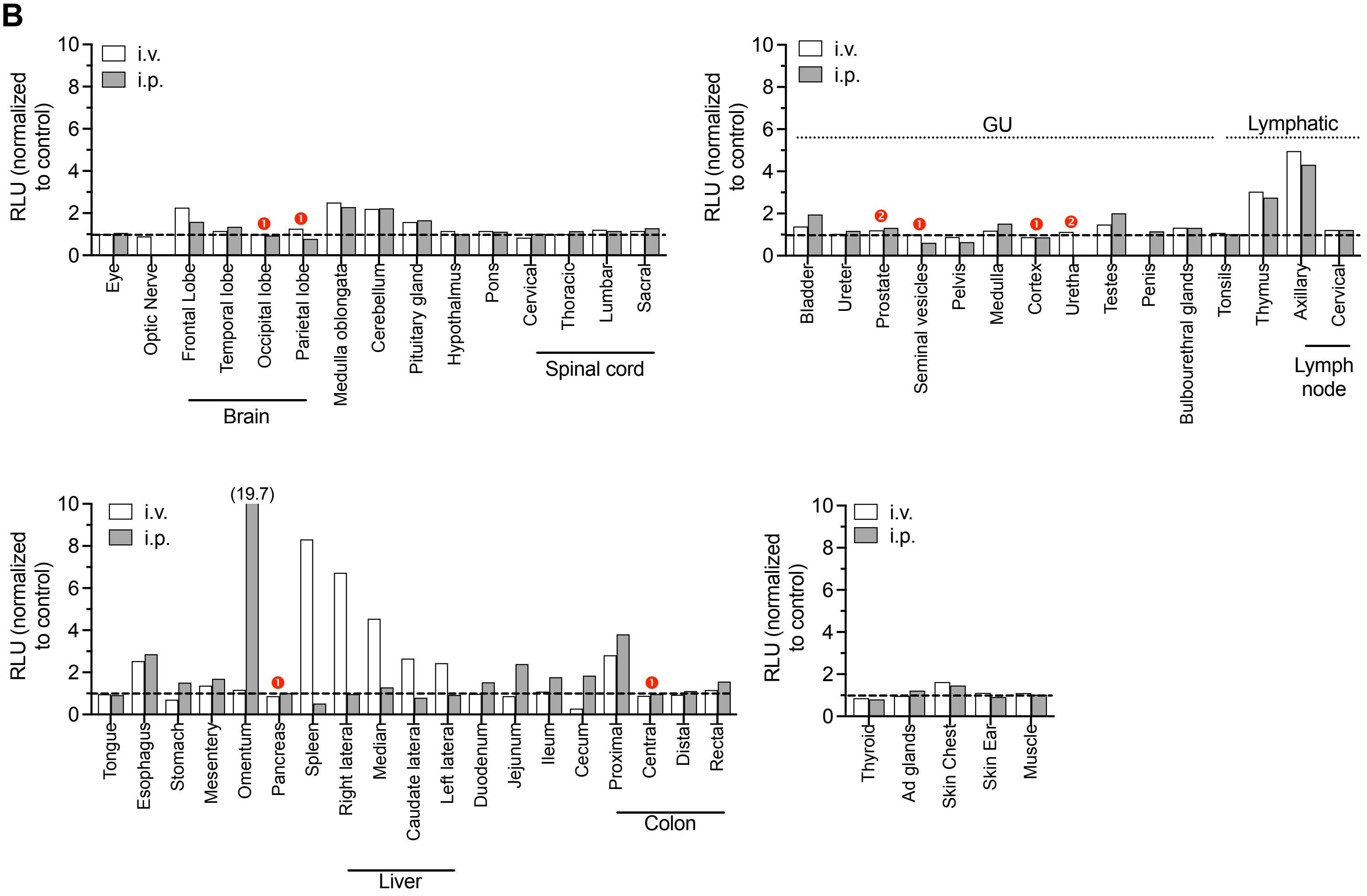

### Supplementary Figure 7

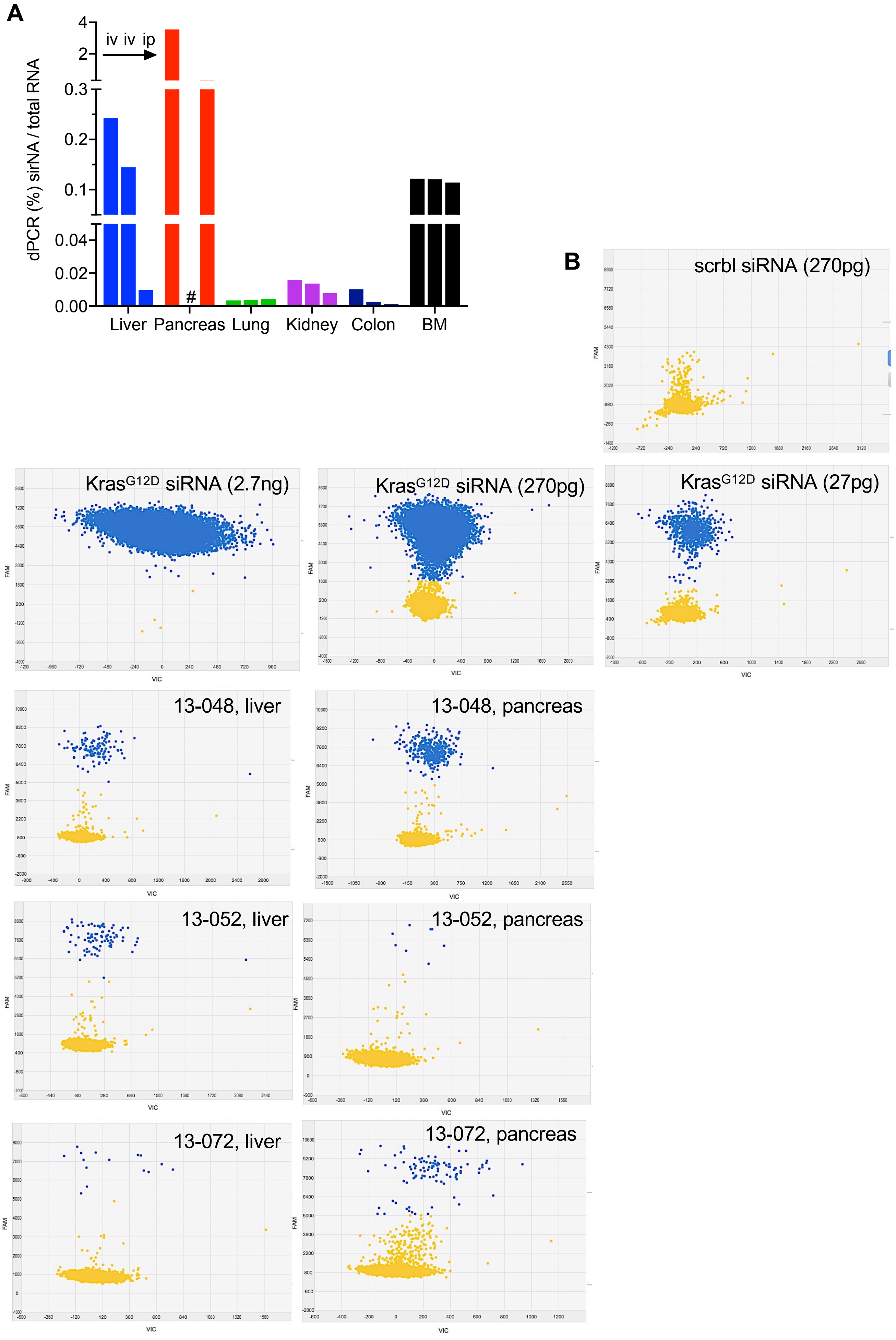

### Supplementary Figure 8

A

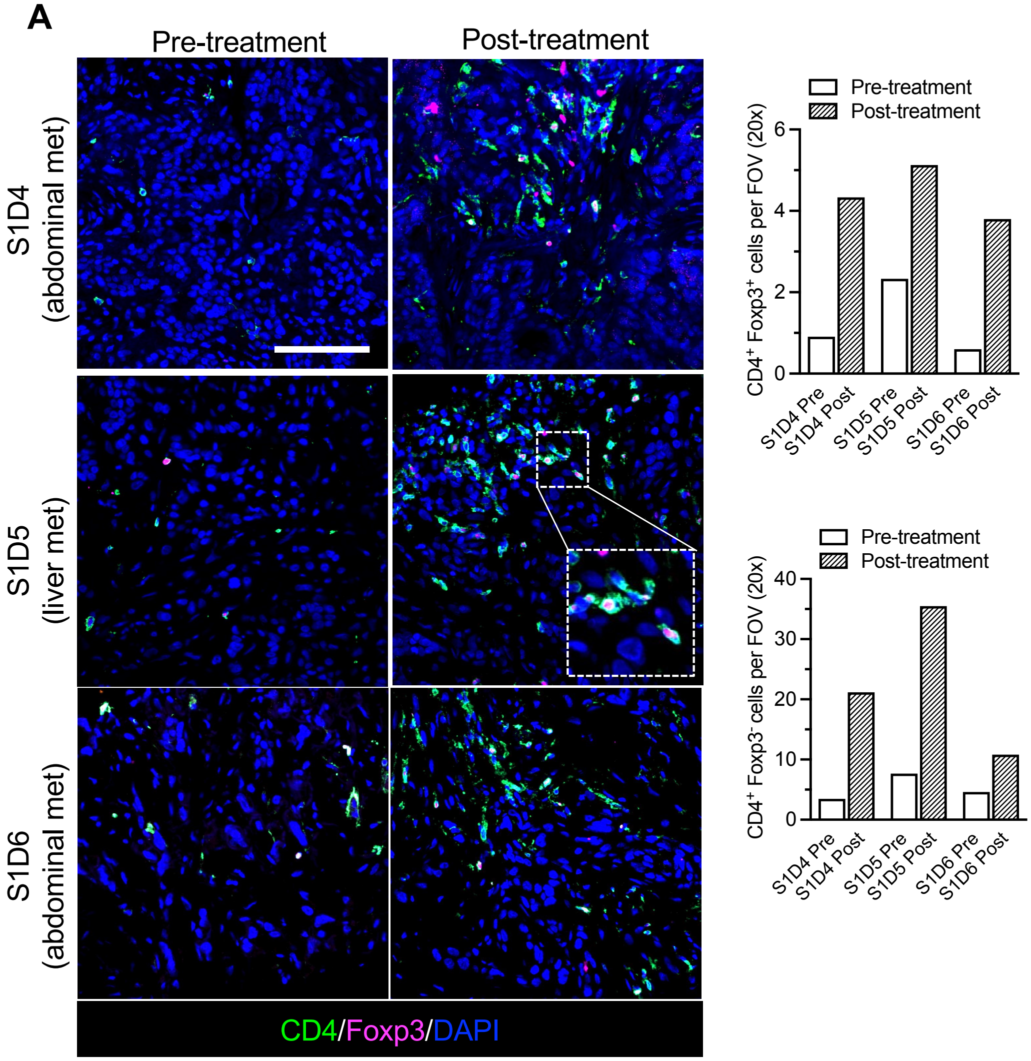


B

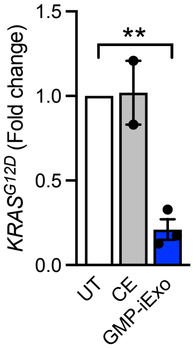

### Supplementary Figure 9

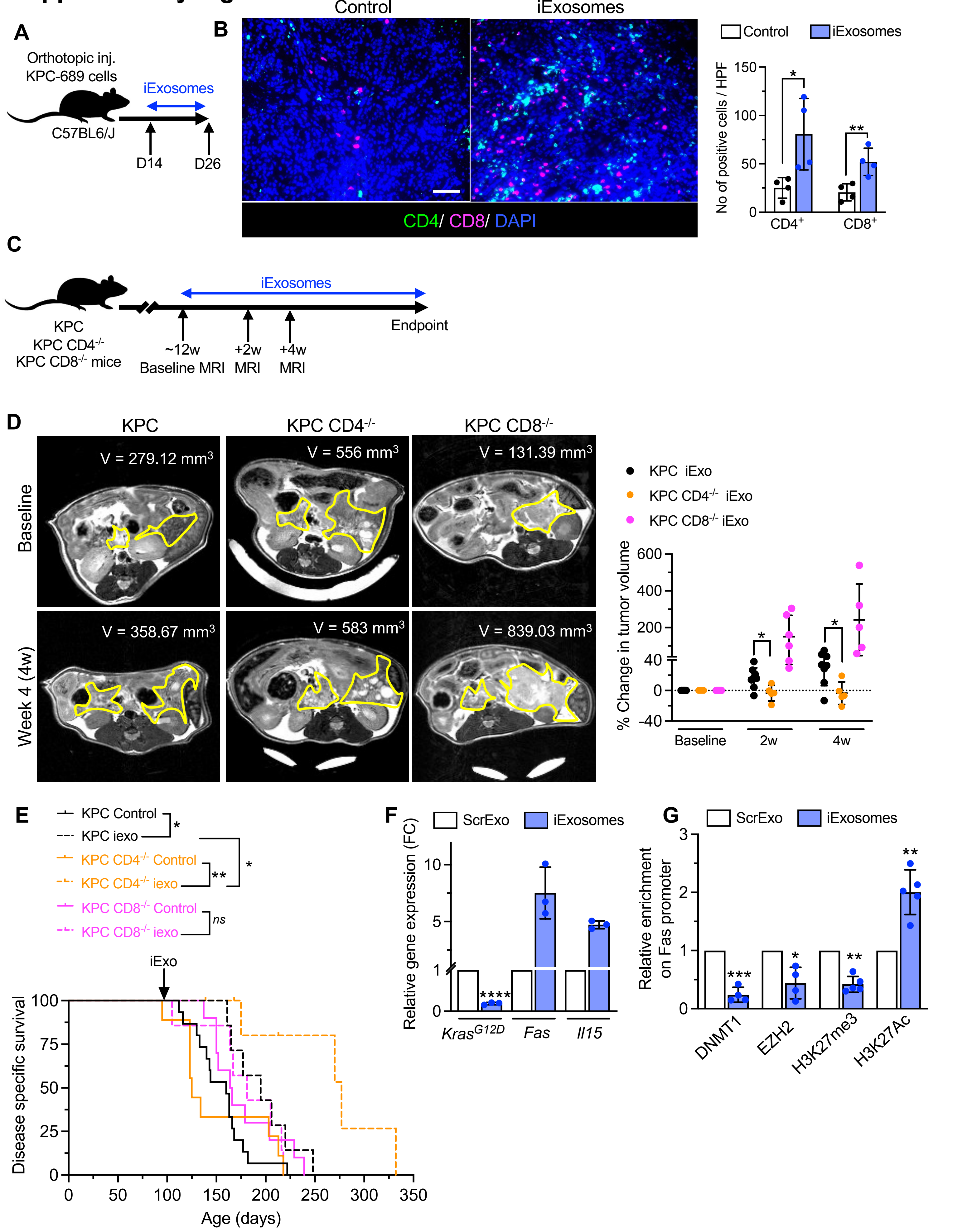

#### Supplementary Figure 10

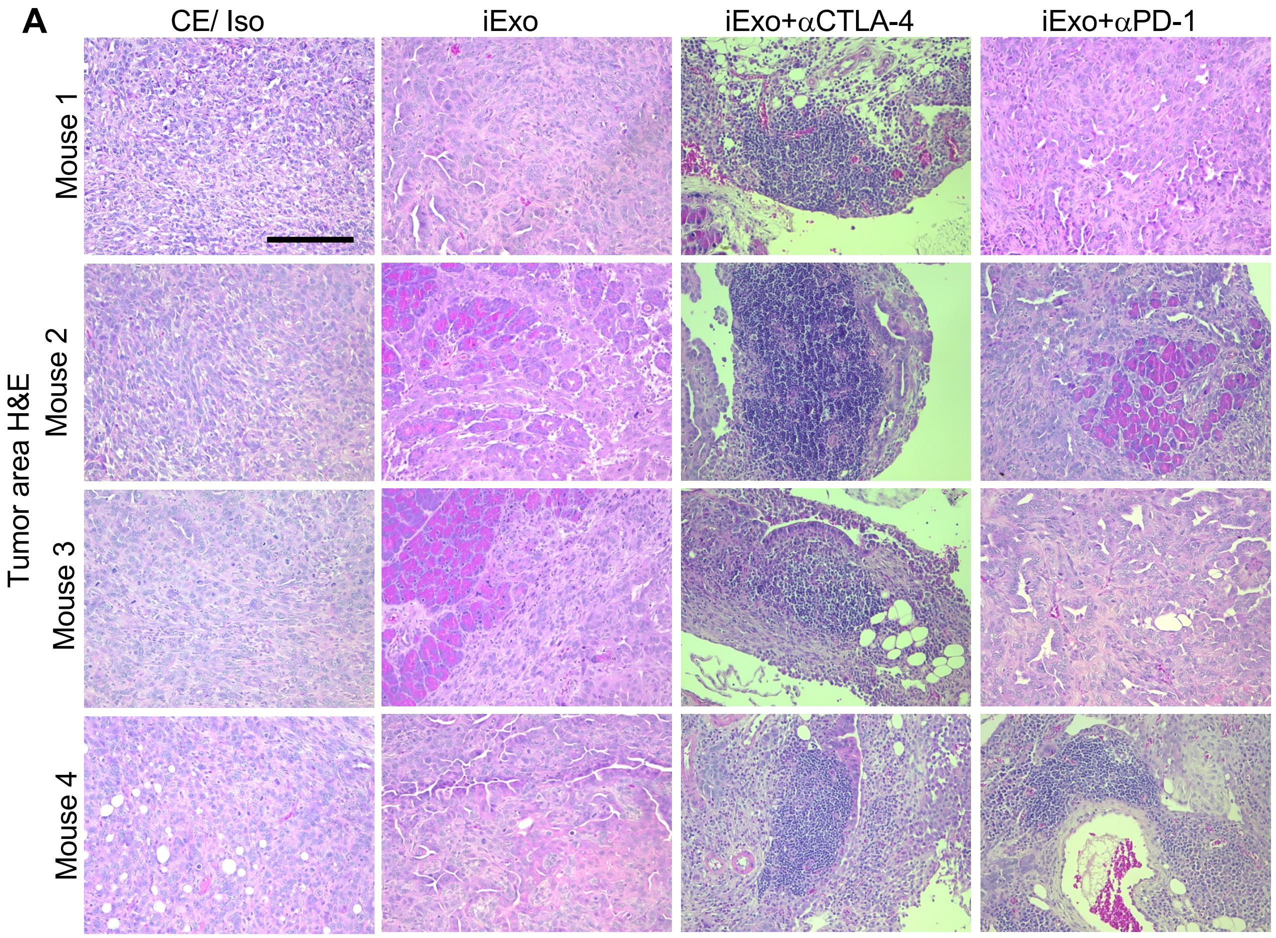


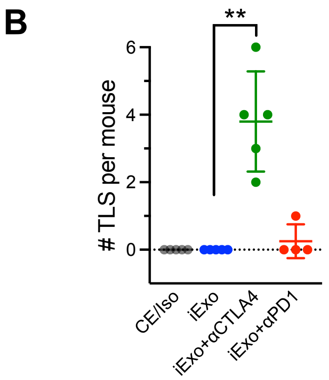
