## Supplementary Table for "KRAS^G12D^-Specific Targeting with Engineered Exosomes Reprograms the Immune Microenvironment to Enable Efficacy of Immune Checkpoint Therapy in PDAC Patients"

**Supplementary Table 1: List of antibodies and TSA reagents**

| Antigen | Primary antibody | | | Secondary antibody | | Opal reagent |
|---|---|---|---|---|---|---|
| | Vendor | Catalog | Concentration | Vendor | Polymer | |
| pERK | Cell signaling technology | 4376T | 1:200 | BioCare | Rabbit-on-Rodent HRP | Opal570 |
| PanCK | Cell signaling technology | 4545S | 1:200 | BioCare | Mouse HRP | Opal690 |
| αSMA | Dako | M0851 | 1:200 | Biocare | Mouse HRP | Opal520 |
| CD8 | Cell signaling technology | 85336S | 1:250 | BioCare | Rabbit-on-Rodent HRP | Opal520 |
| CD8 | Cell signaling technology | 98941s | 1:250 | BioCare | Rabbit-on-Rodent HRP | Opal650 |
| CD4 | Abcam | Ab183685 | 1:400 | BioCare | Rabbit-on-Rodent HRP | Opal520 |
| CK19 | Abcam | Ab52625 | 1:1000 | BioCare | Rabbit-on-Rodent HRP | Opal 520 |
| Fas | Abcam | Ab82419 | 1:100 | BioCare | Rabbit-on-Rodent HRP | Opal 570 |
| CD4 | Abcam | Ab133616 | 1:100 | BioCare | Rabbit on rodent polymer | Opal 520 |
| CD31 (PECAM-1) | Cell signaling technology | D8V9E | 1:100 | BioCare | Rabbit on rodent polymer | Opal 690 |
| FOXP3 | Cell signaling technology | D2W8E | 1:100 | BioCare | Rabbit on rodent polymer | Opal 690 |
| CD19 | Cell signaling technology | D4V4B | 1:100 | BioCare | Rabbit on rodent polymer | Opal 570 |
